# “Maternal and offspring genetic risk score (GRS) analyses of fetal alcohol exposure and ADHD risk in offspring”

**DOI:** 10.1101/2021.03.30.21254492

**Authors:** Elis Haan, Hannah M. Sallis, Eivind Ystrom, Pål Rasmus Njølstad, Ole A. Andreassen, Ted Reichborn-Kjennerud, Marcus R. Munafò, Alexandra Havdahl, Luisa Zuccolo

## Abstract

**Background:** Studies investigating the effects of prenatal alcohol exposure on childhood ADHD symptoms using conventional observational designs have reported inconsistent findings, which may be affected by unmeasured confounding and maternal and fetal ability to metabolise alcohol. We used genetic variants from alcohol metabolising genes (alcohol dehydrogenase (*ADH*) and aldehyde dehydrogenase (*ALDH*)) as proxies for fetal alcohol exposure to investigate their association with offspring ADHD risk around age 7-8.

**Methods:** We used data from three longitudinal pregnancy cohorts: Avon Longitudinal Study of Parents and Children (ALSPAC), Generation R study (GenR) and the Norwegian Mother, Father and Child Cohort study (MoBa). Genetic risk scores (GRS) for alcohol use and metabolism using 36 single nucleotide polymorphisms (SNPs) from *ADH/ALDH* genes were calculated for mothers (N_ALSPAC_=8,196; N_MOBA_=13,614), fathers (N_MOBA_=13,935) and offspring (N_ALSPAC_=8,237; N_MOBA_=14,112; N_GENR_=2,661). Associations between maternal GRS and offspring ADHD risk were tested in the full sample to avoid collider bias. Offspring GRS analyses were stratified by maternal drinking status. Results: The pooled estimate in maternal GRS analyses adjusted for offspring GRS in ALSPAC and MoBa was OR=0.99, 95%CI 0.97-1.02. The pooled estimate in offspring GRS analyses stratified by maternal drinking status across the cohorts was: OR_DRINKING_=0.98, 95%CI 0.94-1.02; OR_NO DRINKING_=0.99, 95%CI 0.97-1.02. These findings remained similar after accounting for maternal genotype data in ALSPAC and maternal and paternal genotype data in MoBa.

**Conclusions:** We did not find evidence for a causal effect of fetal alcohol exposure on ADHD risk in offspring. The results may be affected by low power and outcome assessment.

## 1. Introduction

It is well documented that heavier alcohol consumption during pregnancy can negatively affect fetal development and later neurodevelopmental problems (Patra et al., 2011), but evidence is inconsistent regarding the effects of low prenatal alcohol exposure (PAE) (Mamluk et al., 2017). The harmful neurodevelopmental effects resulting from PAE are collectively defined as fetal alcohol spectrum disorders (FASD), which include fetal alcohol syndrome (FAS), partial FAS, alcohol-related neurodevelopmental disorder (ARND), alcohol-related birth defects (ARBD), and neurobehavioral disorder associated with prenatal alcohol exposure (ND-PAE) (Mattson et al., 2019). However, there is substantial overlap between FASD, attention-deficit hyperactivity disorder (ADHD) and other behavioural impairments (Popova et al., 2016; Weyrauch et al., 2017). A lack of clear diagnostic criteria of FASD makes it difficult to distinguish from other neurodevelopmental disorders (Burd, 2016; Lange et al., 2019). Although ADHD is one of the most common neurodevelopmental disorder diagnoses in FASD, little is known about the role of PAE in causing ADHD symptoms in the general population.

A systematic review by Easey and colleagues suggested that low and moderate alcohol consumption during pregnancy is associated with negative mental health outcomes in children, including anxiety/depression, total behavioural problems and conduct disorder (Easey et al., 2019). However, another recent systematic review and meta-analysis focusing specifically on offspring ADHD found little evidence to suggest an increased risk of ADHD symptoms in children whose mothers consumed moderate amounts of alcohol (up to 70 grams a week) during pregnancy (Porter et al., 2019). In contrast, low PAE was found to have a protective effect on ADHD symptoms in some earlier studies (Kelly et al., 2013; Niclasen et al., 2014). However, it is possible that these associations are due to genetic confounding or confounding by social factors, as both studies found that women who drank low or moderate levels during pregnancy had a higher socio-economic position, which is associated with ADHD risk in the offspring. Furthermore, studies based on siblings discordant on PAE that account for shared genetics, can strengthen evidence for causality. In these studies, PAE showed a minor residual effect on internalizing problems up to age 3 (Lund et al., 2019) and ADHD symptoms at 5 years (Eilertsen et al., 2017). Most of the effect was accounted for by maternal factors shared by siblings. In these studies, bias because of unmeasured confounding and non-shared environmental factors cannot be ruled out.

One potential approach to overcome limitations due to unmeasured confounding is to use genetic variants predictive of alcohol consumption or directly involved in alcohol metabolism to disentangle effects of PAE on child outcomes. Using a Mendelian Randomization (MR) design can also help to investigate causality, as the genetic variants used as a proxy for the exposure are generally less affected by confounding factors than the self-reported exposure measurements used in more conventional epidemiological analyses (Davey Smith and Hemani, 2014). If we observe evidence of an association between the genetic proxy and the outcome, this provides stronger support for a causal influence of the exposure (prenatal alcohol consumption) on our outcome (offspring ADHD risk) (Davey Smith and Ebrahim, 2003).

Our recent report using a combination of negative control and polygenic risk score (PRS) analyses, did not find evidence for a causal effect of PAE on offspring ADHD risk (Haan et al., 2021). In this previous study, the alcohol PRS was derived using SNPs identified from the latest GWAS on alcohol consumption per week (Liu et al., 2019). However, the alcohol PRS was based on a discovery sample of general population individuals, who on average consume more alcohol than pregnant women. Moreover, there is evidence that harmful effects of alcohol exposure are also affected by maternal and fetal metabolic capacity which can lead to different fetal alcohol levels if pregnant women drink alcohol (Burd et al., 2012). Therefore, analyses based on the PRS of alcohol consumption per week could miss biologically important effects related to individual differences in the ability to metabolise alcohol. A large body of research has shown that ethanol metabolism is affected by a group of alcohol dehydrogenase (*ADH: ADH1A, ADH1B, ADH1C, ADH4, ADH5, ADH6, ADH7*) and aldehyde dehydrogenase (*ALDH: ALDH1A1* and *ALDH2*) genes, and variants in these genes impact how quickly alcohol is metabolised, the amount of alcohol consumed and individual effects of alcohol consumption (Birley et al., 2009; Birley et al., 2008; Edenberg and McClintick, 2018). Furthermore, genetic variants of ADH genes are also expressed in early fetal development. After maternal alcohol intake, fetal blood alcohol concentration is nearly equivalent to maternal alcohol levels (Burd et al., 2012) and although the fetus can metabolise some alcohol, the majority of alcohol metabolism acts through maternal metabolic pathways (Burd et al., 2012). The importance of fetal ADH genetic variants is evidenced by previous studies using the ALSPAC sample which have shown that four child *ADH* genetic variants were associated with lower IQ at age 8 and early onset conduct problems in children whose mothers drank during pregnancy (Lewis et al., 2012; Murray et al., 2016). However, these studies used only limited number of *ADH* genetic variants selected based on the association with the outcome, and it is still unknown how these four genetic variants change metabolic activity.

It remains unclear whether PAE has a causal effect on offspring ADHD risk, through modulations of maternal and fetal alcohol metabolism. In this study, we use genetic variants in *ADH/ALDH* as proxies for fetal alcohol exposure and investigate their association with offspring ADHD risk, and separately with hyperactive-impulsive and inattention symptom domains, using data from three large European birth cohorts (Figure 1).

**Figure 1.**
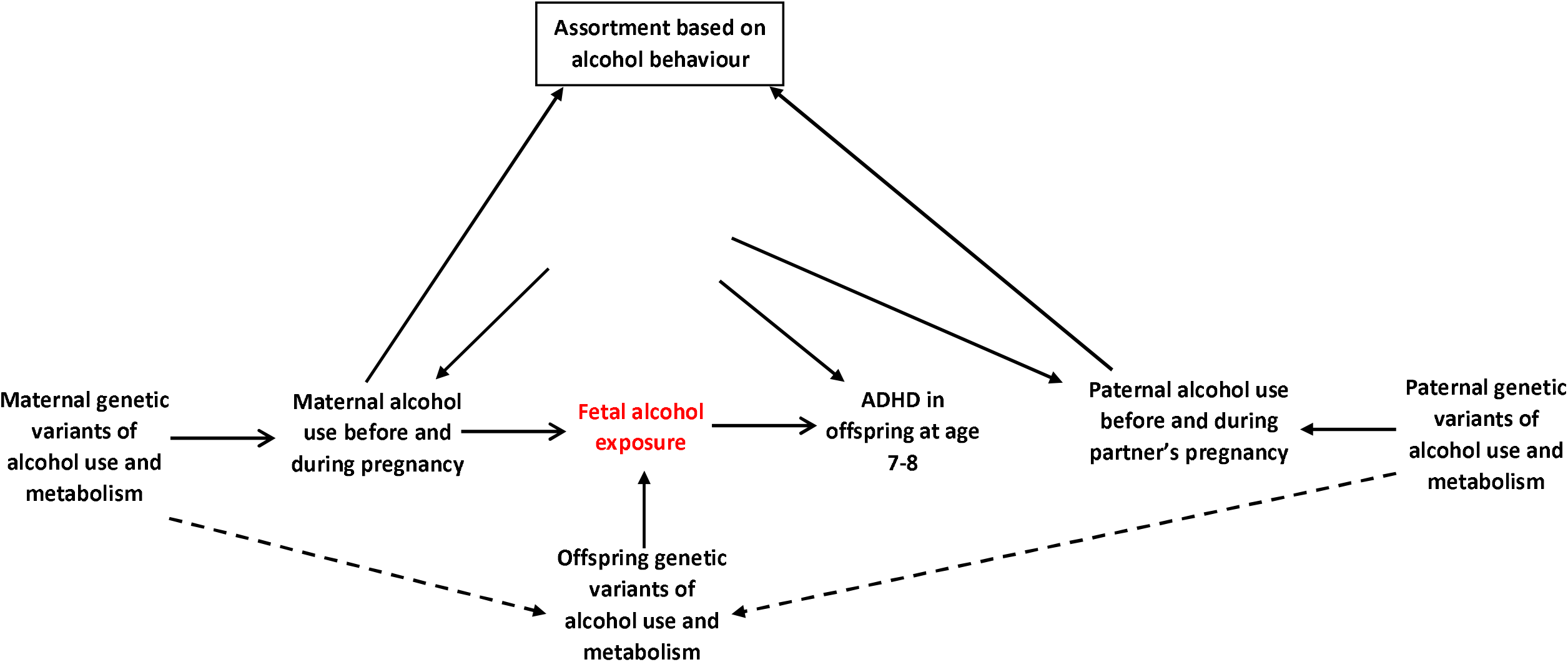
Study design. Note: Maternal and Offspring genetic variants of alcohol use and metabolism were used as proxies for fetal alcohol exposure (the exposure of interest, unmeasured) to investigate associations with ADHD symptoms in offspring around age 7 years (outcome). Dashed arrows represent genetic correlation as a child inherits 50% of its genetic make-up from mother and father. In this study we assume that mate could be affected by alcohol behaviour. Adjustment for paternal genetic data will help to overcome potential bias due to the assortative mating behaviour.

## 2. Methods

### 2.1. Study populations

We used data from three European prospective longitudinal birth cohorts: the Avon Longitudinal Study of Parents and Children (ALSPAC), Generation R (GenR) and the Norwegian Mother, Father and Child Cohort Study (MoBa). ALSPAC is a prospective longitudinal cohort study that recruited 14,541 pregnant women resident in Avon, UK with expected dates of delivery between 1st April 1991 and 31st December 1992 (Boyd et al., 2013; Fraser et al., 2013; Northstone et al., 2019). Generation R (GenR) is a population-based prospective cohort study in Rotterdam in the Netherlands that recruited 9,778 pregnant women expected to give birth between April 2002 and January 2006 (Kooijman et al., 2016). MoBa is a population-based pregnancy cohort study where participants were recruited from all over Norway between 1999 and 2008. The cohort now includes 114,500 children, 95,200 mothers and 75,200 fathers (Magnus et al., 2016). More details are shown in the Supplementary Material.

### 2.2. Availability of genome-wide data

In ALSPAC, genome-wide data are available for 8,237 children and 8,196 mothers. In GenR, genetic data are available for 2,661 children from European ancestry, but maternal genetic data was not available at the time of analyses. In MoBa, genetic data are currently available for 14,112 children, 13,614 mothers and 13,935 fathers. Detailed information about the genotyping is presented in the Supplementary Material.

### 2.3. Exposures

In ALSPAC, at 18 weeks gestation mothers were asked about their average amount and frequency of alcohol consumption a) during the first trimester, b) in the previous 2 weeks, c) at the time when they first felt the baby move. At 32 weeks gestation mothers were asked about their average weekday and weekend alcohol consumption.

In GenR, in the first questionnaire (<18 weeks) mothers were asked whether they drank any alcohol in the first 3 months of pregnancy. In mid (18-25 weeks) and late pregnancy (>25 weeks) mothers were asked if they drank alcohol in the past 2 months and mothers who reported drinking were asked to classify their average alcohol consumption.

In MoBa, at 15 and 30 weeks gestation mothers were asked how often and how many units of alcohol they consumed in the current pregnancy. 6 months after their child’s birth, mothers were asked about their alcohol consumption (amount and frequency) during the last 3 months of their pregnancy.

Mothers who reported alcohol consumption at any point during the pregnancy were classified as drinkers. More details are presented in Supplementary Table S1.

### 2.4. Outcome

ADHD was measured around age 7-8 years in each cohort. As in our previous study (Haan et al., 2021) we wanted to explore whether we observe differences in ADHD hyperactive-impulsive and inattention symptom domains, if ADHD symptoms were reported by mother or teacher, and whether results depended on the questionnaire used for ADHD assessment.

As the continuous score of ADHD symptoms was either zero-inflated or skewed, a binary variable was derived for total ADHD, hyperactive-impulsive and inattention symptoms using the 85^th^ percentile as a threshold to indicate presence of high level symptoms (Achenbach and Rescorla, 2001; Aylward and Stancin, 2008). Up to 4 missing items were allowed depending on the total number of items in the questionnaire. More details can be found in Supplementary Table S2.

#### 2.4.1. Primary outcome measure

The psychometric scales used for the main outcome measure were: maternal report of the Development And Well-Being Assessment (DAWBA) questionnaire in ALSPAC; maternal report of the revised Conner’s Parent Rating Scale (CPRS-R) in GenR; and maternal report of the Disruptive Behaviour Disorders scale (RS-DBD) in MoBa.

#### 2.4.2. Secondary outcome measures

We additionally included teacher report of the DAWBA questionnaire and maternal and teacher report of the Strength and Difficulties Questionnaire (SDQ) hyperactivity subscale in ALSPAC, and maternal and teacher report of the Child Behaviour Checklist (CBCL) attention problems subscale in GenR.

### 2.5. Genetic variants

Based on previous research we identified genes responsible for alcohol metabolism that were expressed in liver and brain using Expression Atlas (Kapushesky et al., 2010; Papatheodorou et al., 2020). Using this online tool, we identified *ADH1A, ADH1B, ADH4, ADH5, ADH6, ALDH2, ALDH1A1* and *ALDH1B1* that were expressed in adults and fetus, and additionally *ADH1C* and *ADH7* genes which were only expressed in adults. All these genes are located in chromosomes 4, 9 and 12. In total 869 single nucleotide polymorphisms (SNPs) from these genes were identified in ALSPAC (Supplementary Figures S1-S3). Given the high linkage disequilibrium of these SNPs, we used a clumping procedure (R^2^<0.01) to identify independent SNPs. 1000 Genomes was used as a reference panel for clumping. After clumping, we identified 36 independent SNPs (Supplementary Table S3).

### 2.6. Harmonisation

Before calculating genetic risk scores (GRS), SNPs were aligned so that the effect alleles were positively associated with alcohol consumption in the discovery sample. These effect estimates were taken from the summary statistics of the latest GWAS on alcohol consumption per week (Liu et al., 2019). The harmonisation procedure in each cohort is shown in Supplementary Tables S4-S6. All SNPs identified in ALSPAC were also available in GenR. In MoBa, genetic imputation was conducted using the Haplotype Reference Consortium (HRC) as a reference panel, and 9 SNPs identified in ALSPAC were unavailable in MoBa. We identified proxy for these SNPs using Single Nucleotide Polymorphisms Annotator (SNiPA) (Arnold et al., 2015) (Supplementary Table S6).

### 2.7. Genetic risk scores

As it is not possible to directly measure the effect of each SNP on fetal exposure to alcohol, and therefore to calculate weights, we derived unweighted offspring and parental GRS using PLINK 1.9. Generation R data only allowed computation of offspring GRS, ALSPAC allowed computation of offspring and maternal GRS, and MoBa allowed computation of offspring, maternal and paternal GRS.

### 2.8. Statistical analyses

All analyses were performed using Stata (v15: ALSPAC, GenR; v16: MoBa) (StataCorp, 2017, 2019) and restricted to unrelated individuals of European ancestry. Analyses were performed as described in our pre-registered protocol (Haan et al., 2019). Associations between maternal and offspring GRS and ADHD risk in offspring were tested using logistic regression. All analyses were adjusted for 10 ancestry informed principal components and in MoBa additionally for birth year and genotyping batch. Thanks to the harmonisation procedure described above, estimates of association between GRS and ADHD risk can be interpreted as odds ratios (ORs) per additional alcohol-increasing GRS allele – a positive OR likely indicates a detrimental effect of fetal alcohol exposure on ADHD risk.

### 2.8.1. Primary analysis

We carried out analyses using both maternal and offspring GRS as proxies for increased fetal alcohol exposure. Maternal analyses were performed using three models: 1) maternal GRS; 2) maternal GRS adjusted for offspring GRS; 3) maternal GRS adjusted for offspring and paternal GRS. These analyses were performed in the full sample without stratifying based on maternal drinking status, because this could induce collider bias as genetic propensities of women who drink during pregnancy may differ from those women who abstain, thus introducing a spurious association between the GRS and confounders (Supplementary Figure S4a). Three models were used also in offspring GRS analyses: 1) offspring GRS; 2) offspring GRS adjusted for maternal GRS; 3) offspring GRS adjusted for maternal and paternal GRS. These analyses were stratified by maternal drinking status during pregnancy, since collider bias is not be an issue when we are interested in fetal alcohol exposure (Supplementary Figure S4b).

Analyses were performed separately in each cohort and then meta-analysed across the cohorts using a random effects model which accounts for variability in the exposure and outcome assessment across the cohorts. ALSPAC and MoBa results from model 2 (maternal GRS adjusted for offspring GRS) were pooled in meta-analyses, while all three cohorts contributed to meta-analyses for the offspring GRS model 1 results.

#### 2.8.2. Sensitivity analyses

We also checked the influence of each individual SNP on the outcome using a leave-one-out approach. We created 36 additional GRS for mothers and offspring excluding one SNP at a time. Given that there was no deviation in effect estimates when leaving out individual SNPs (Supplementary Figures S5-15), we used the GRS including all 36 SNPs in our analyses. We also tested the associations between maternal GRS and potential confounders.

#### 2.8.3. Replication analyses of previous ALSPAC studies

We also created a GRS including four offspring ADH genetic variants (*ADH1A*: rs975833, rs2866151; *ADH1B*: rs4147536; *ADH7* rs284779) found to be associated with neurodevelopmental outcomes in previous ALSPAC studies (Lewis et al., 2012; Murray et al., 2016).

## 3. Results

An overview of study sample characteristics is shown in Table 1. In all the cohorts, mothers who reported drinking were older and better educated, compared to non-drinking mothers.

**Table 1.** Study sample characteristics.

|  | ALSPAC |  | GenR |  | MoBa |  |
| --- | --- | --- | --- | --- | --- | --- |
|  | Maternal alcohol consumption during pregnancy |  |  |  |  |  |
|  | No<br>N=1,125 | Yes<br>N=2,573 | No<br>N=1,004 | Yes<br>N=1,659 | No<br>N=6,536 | Yes<br>N=1,357 |
| Confounders |  |  |  |  |  |  |
| Mother's age in years<br>(mean and SD) | 28 (4.5) | 30 (4.3) | 30 (4.8) | 32 (4.1) | 30 (4.3) | 32 (3.9) |
| Mother's education |  |  |  |  |  |  |
| Primary | 14% | 8% | 10% | 2% | 1% | 1% |
| Secondary | 71% | 69% | 49% | 28% | 27% | 19% |
| Higher | 15% | 23% | 41% | 70% | 72% | 80% |
| Financial difficulties* |  |  |  |  |  |  |
| No | 42% | 40% | 80% | 90% | 87% | 86% |
| Yes | 58% | 60% | 20% | 10% | 13% | 14% |
| Marital status |  |  |  |  |  |  |
| Married | 84% | 85% | 92% | 92% | 98.5% | 98% |
| Single/not married | 16% | 15% | 8% | 8% | 1.5% | 2% |
| Depression symptoms** | 9% | 10% | 8% | 5% | 5% | 6% |
| Anxiety symptoms | 12% | 14% | 10% | 6% |  |  |
| Mothers ADHD symptoms |  |  |  |  | 2% | 3% |
| Parity |  |  |  |  |  |  |
| 1st | 53% | 45% | 57% | 61% | 51% | 40% |
| 2nd | 33% | 37% | 30% | 30% | 33% | 39% |
| 3rd+ | 14% | 18% | 13% | 9% | 16% | 21% |
| Mother smoked during pregnancy | 15% | 17% | 17% | 25% | 5% | 7% |
| Child ADHD symptoms above the 85th percentile threshold | 13% | 14% | 13% | 15% | 12% | 12% |

\* In ALSPAC, financial difficulties were measured with 5 items questionnaire: 1) Difficulty in affording food; 2) Difficulty in affording clothing; 3) Difficulty in affording heating 4) Difficulty in affording accommodation 5) Difficulty in affording things for baby. In GenR, financial difficulties were assessed with single item question: Difficulty in paying food, rent, bills and suchlike. In MoBa, financial difficulties were assessed with single item question: Have you experienced financial problems?
\*\*In MoBa depression and anxiety problems were assessed together

### 3.1. Maternal GRS analysis

The pooled estimate for the association of maternal GRS with high risk of maternal reported ADHD symptoms in offspring (model 2 - adjusted for offspring GRS) did not show evidence for an association (OR_ADHD_=0.99, 95%CI 0.97-1.02; OR_HYP_=0.99, 95%CI 0.95-1.03; OR_INA_=1.00, 95%CI 0.97-1.02) (Figure 2). In MoBa we found weak evidence of a negative association between maternal GRS and high risk of maternal reported offspring symptoms in the hyperactivity domain only, in model 1 (maternal GRS) and model 3 (adjusted for offspring and paternal GRS) (Figure 3, Supplementary Table S7). This finding was not replicated in ALSPAC using either maternal or teacher report (Figure 3, Supplementary Figure S16, Supplementary Tables S8 & S9).

**Figure 2.**
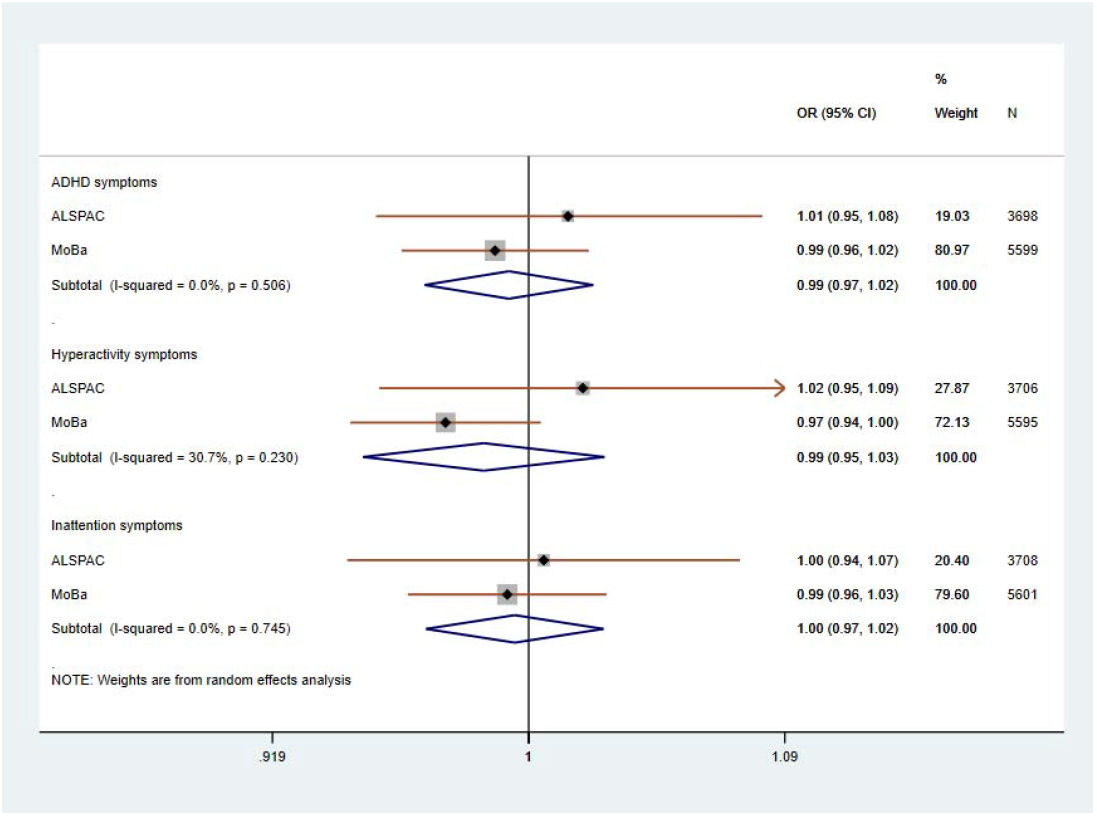
Meta-analysis of maternal alcohol GRS on high risk of maternal reported offspring ADHD symptoms in ALSPAC and MoBa. Note: Model 2 – maternal GRS adjusted for offspring GRS and 10 ancestry principal components

**Figure 3.**
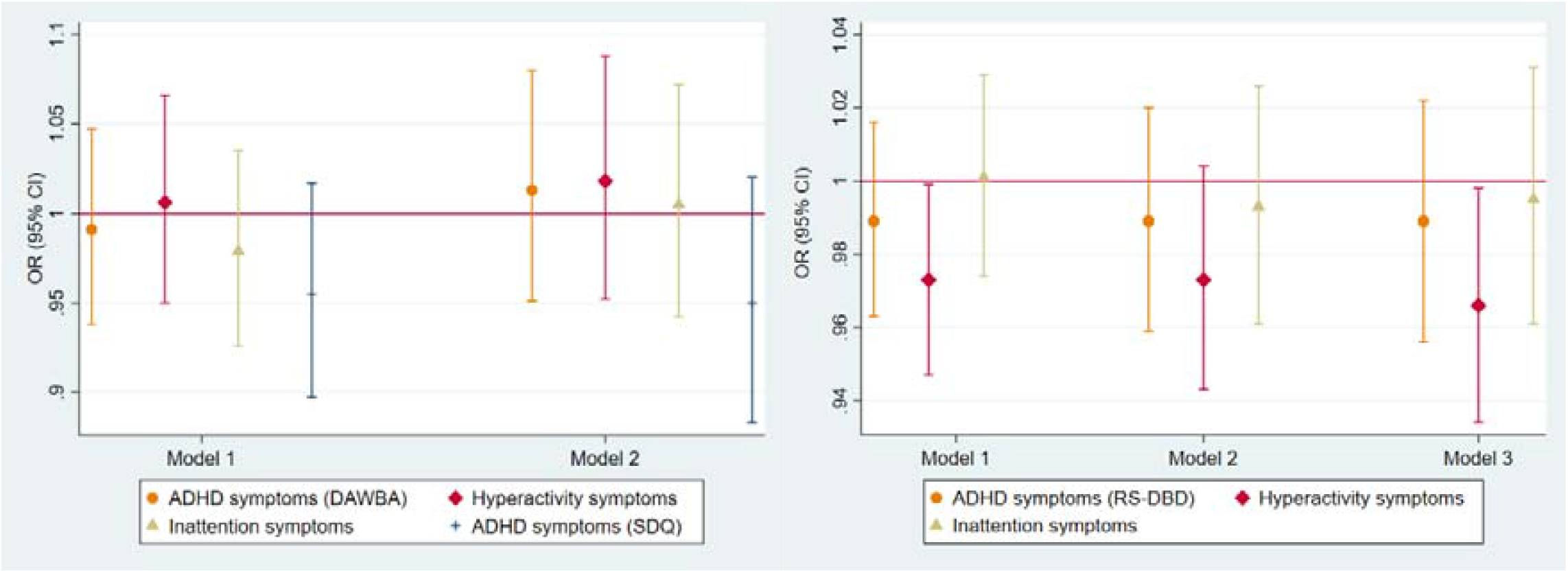
Associations between maternal alcohol GRS and high risk of maternal reported offspring ADHD symptoms in ALSPAC and MoBa. Note: Model 1 – only maternal GRS; Model 2 – maternal GRS adjusted for offspring GRS; Model 3 - maternal GRS adjusted for offspring and paternal GRS; all analyses are adjusted for 10 ancestry principal components. In MoBa, also for birth year and genotyping batch; Development And Well-Being Assessment (DAWBA); Strength and Difficulties Questionnaire (SDQ) – secondary measure; Disruptive Behaviour Disorders scale (RS-DBD)

### 3.2. Offspring GRS analysis

The pooled estimate of the association between offspring GRS and maternal reported ADHD risk in offspring using model 1 (offspring GRS) found no strong evidence of an effect. This remained consistent regardless of drinking status during pregnancy (Drinking: OR_ADHD_=0.98, 95%CI 0.94-1.02; OR_HYP_=0.99, 95%CI 0.95-1.03; OR_INA_=0.98, 95%CI 0.94-1.02; No drinking: OR_ADHD_=0.99, 95%CI 0.97-1.02; OR_HYP_=0.99, 95%CI 0.96-1.01; OR_INA_=0.99, 95%CI 0.91-1.09) (Figure 4). These results did not change after adjusting for maternal GRS (model 2) or maternal and paternal GRS (model 3) in ALSPAC and MoBa (Figure 5 and 6, Supplementary Tables S10-S13). Similarly, when using secondary outcome measures in GenR and ALSPAC, we found no strong evidence of an association between offspring GRS and ADHD risk in offspring (Figure 7, Supplementary Figure S17, Supplementary Tables S14-S16).

**Figure 4.**
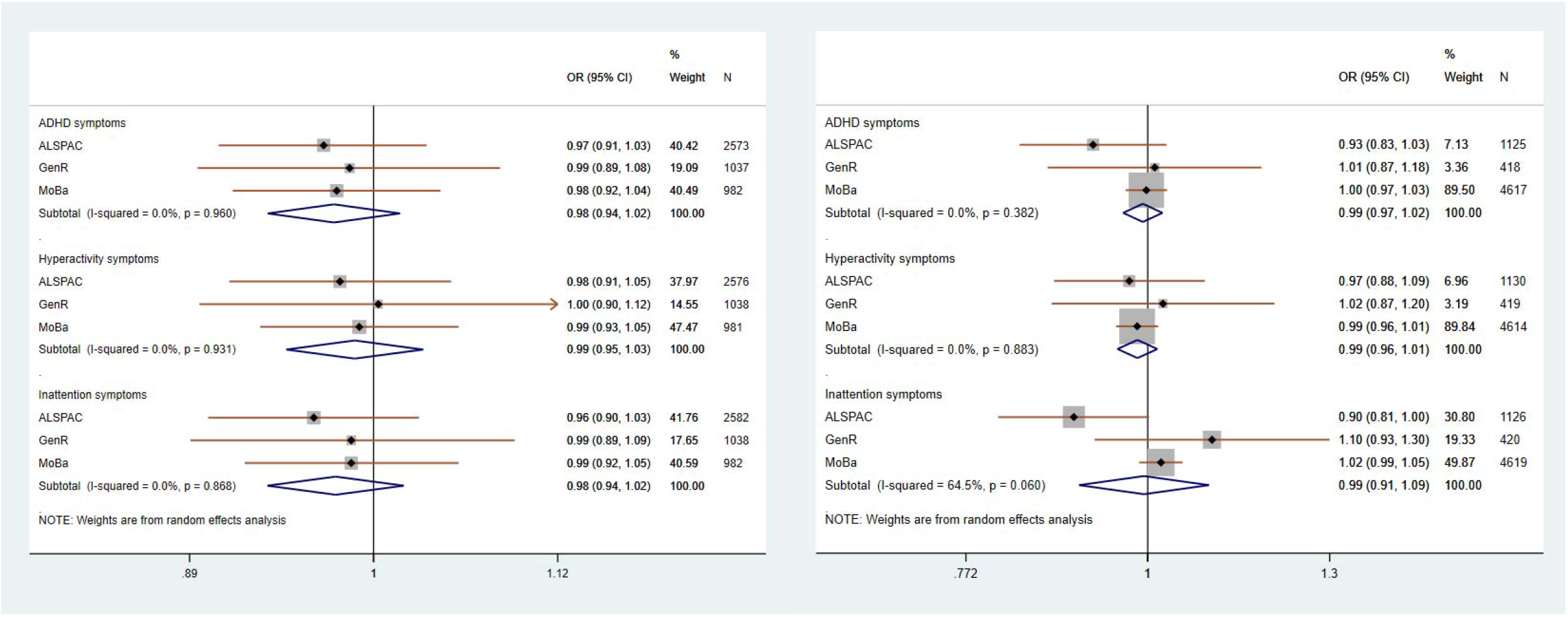
Meta-analysis of offspring GRS on high risk of maternal reported offspring ADHD symptoms – ALSPAC, Generation R and MoBa. Note: Model 1 – only offspring GRS and adjusted for 10 ancestry principal components

**Figure 5.**
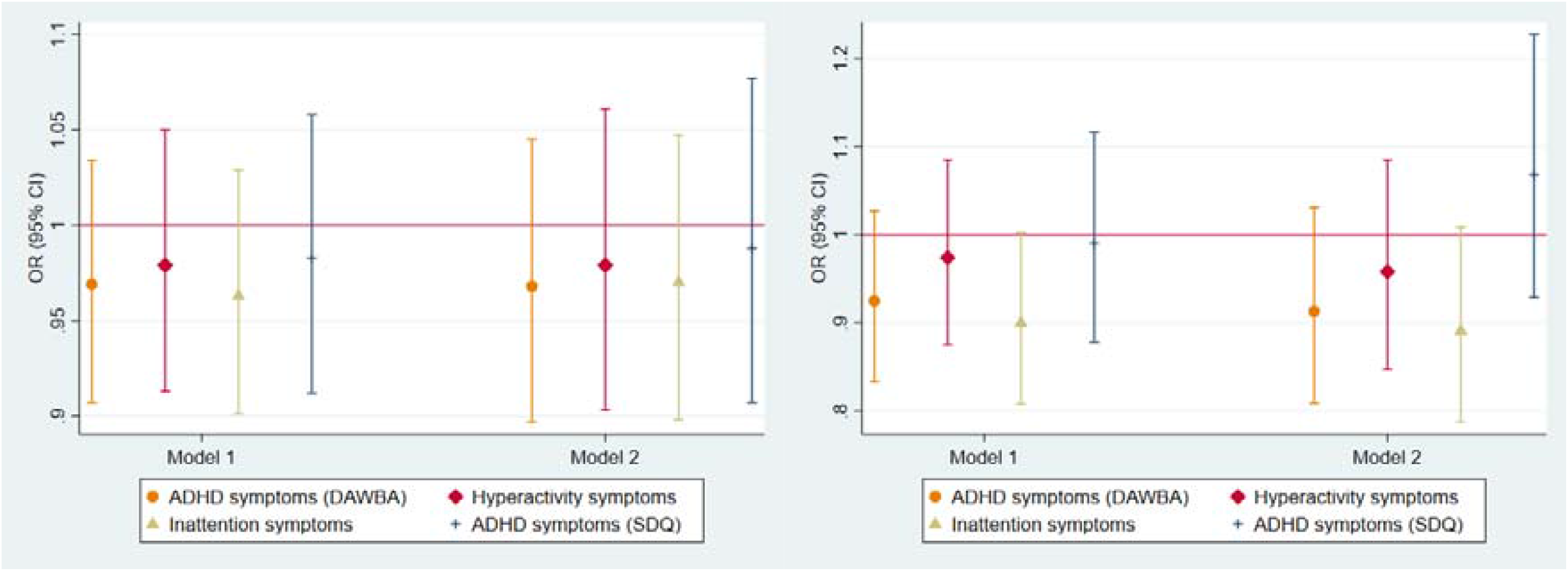
Associations between offspring GRS and high risk of maternal reported offspring ADHD symptoms in ALSPAC stratified by maternal drinking status. Note: Model 1 – only offspring GRS; Model 2 – offspring GRS adjusted for maternal GRS; all analyses adjusted for 10 ancestry principal components; Development And Well-Being Assessment (DAWBA); Strength and Difficulties Questionnaire (SDQ) – secondary measure

**Figure 6.**
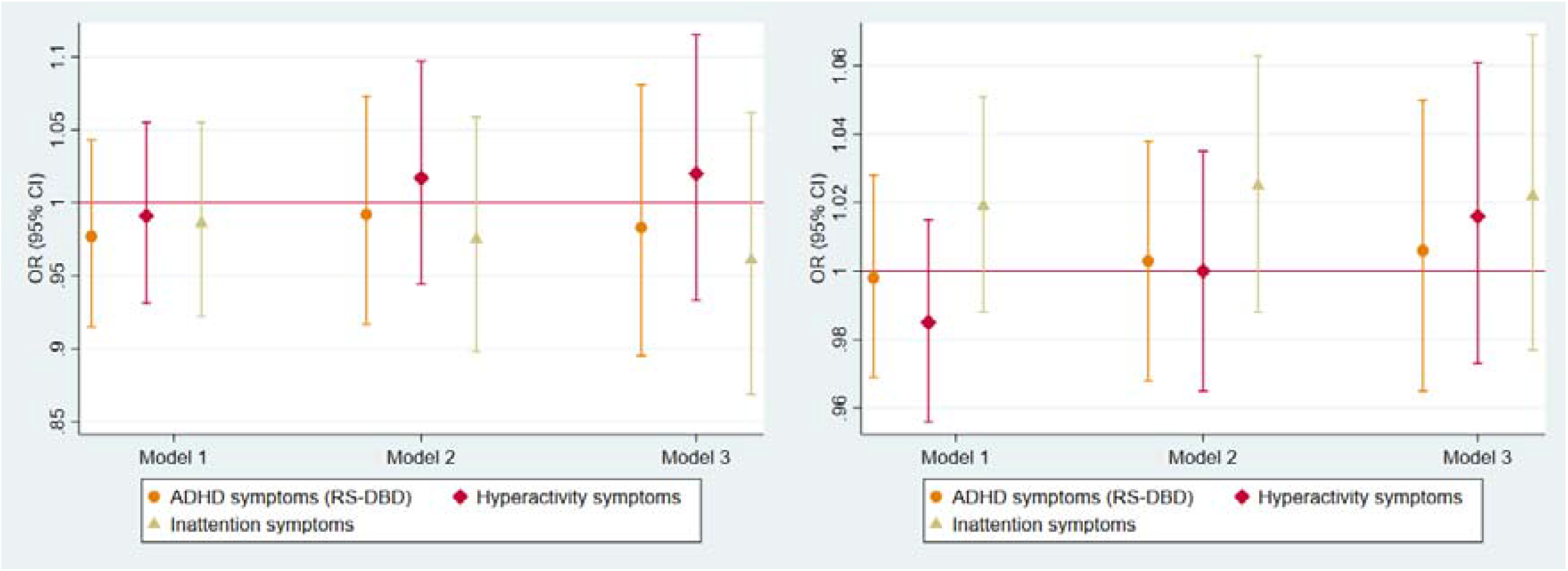
Associations between offspring GRS and high risk of maternal reported offspring ADHD symptoms in MoBa stratified by maternal drinking status. Note: Model 1 – only offspring GRS; Model 2 – offspring GRS adjusted for maternal GRS; Model 3 – offspring GRS adjusted for maternal and paternal GRS; all analyses are adjusted for 10 ancestry principal components, birth year and genotyping batch; Disruptive Behaviour Disorders scale (RS-DBD)

**Figure 7.**
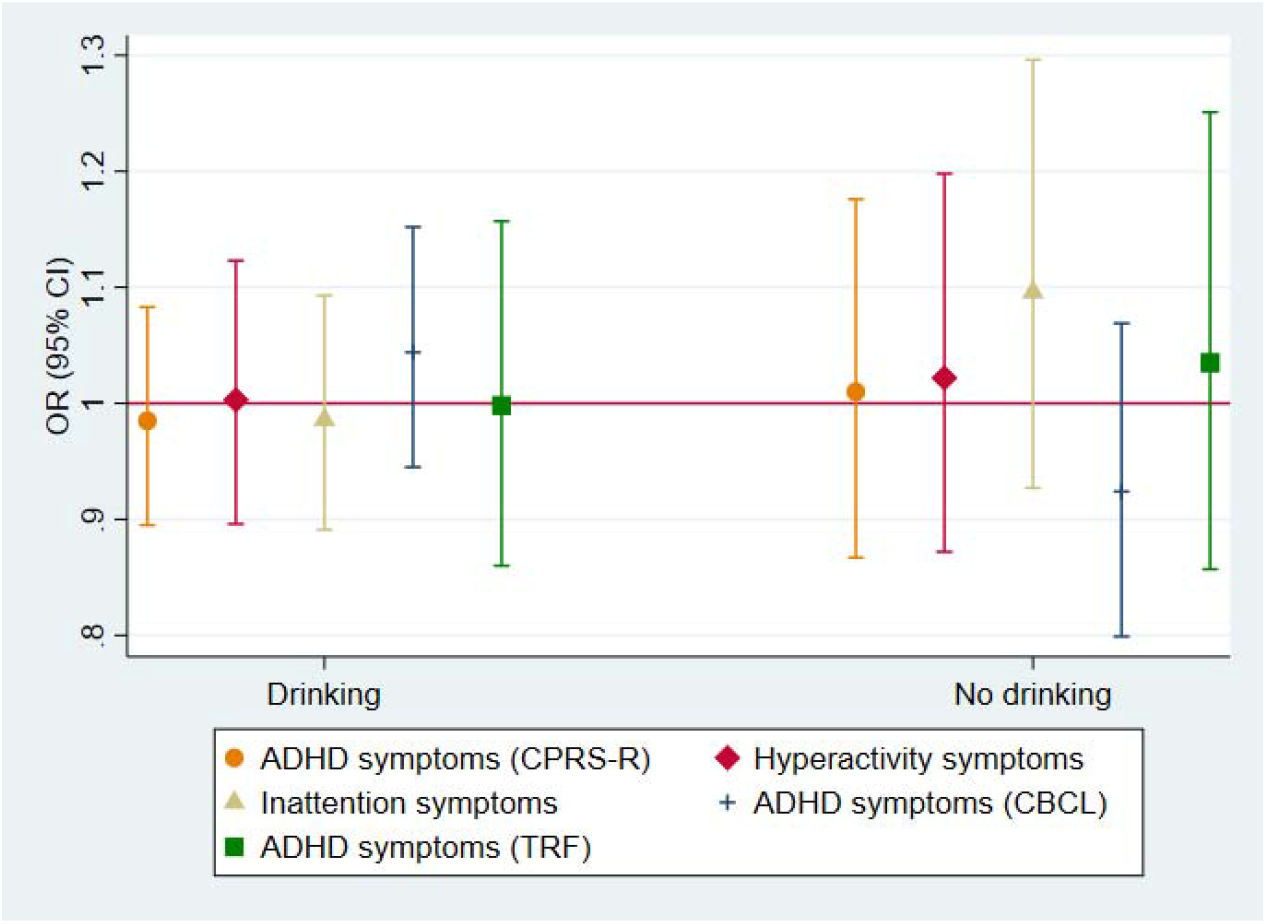
Associations between offspring GRS and high risk of maternal and teacher reported offspring ADHD symptoms in GenR stratified by maternal drinking status. Note: Model 1 – only offspring GRS adjusted for 10 ancestry principal components; Revised Conner’s Parent Rating Scale (CPRS-R); Child Behaviour Checklist (CBCL); Teacher Report Form (TRF); CBCL and TRF are secondary measures

### 3.3. Sensitivity analysis

Sensitivity analyses testing the associations between maternal GRS and confounders found an association with higher likelihood not being married in MoBa, but no associations were observed in ALSPAC (Supplementary Table S17-S18).

### 3.4. Replication analysis of previous ALSPAC studies

Replication analysis using offspring GRS with 4 ADH SNPs were consistent with the main analysis, and provided little evidence for an association between fetal alcohol exposure and high risk of maternal or teacher reported ADHD symptoms in offspring (Supplementary Figures S18-S21 and Supplementary Tables S19-S25).

## 4. Discussion

We used maternal and offspring variants in the *ADH* and *ALDH* genes linked to alcohol use and metabolism to investigate whether there is a causal effect of fetal alcohol exposure on childhood ADHD risk. Consistently with previous work, we did not find evidence for a causal effect (Haan et al., 2021). We observed only weak evidence in one cohort (MoBa) of a negative association between maternal GRS and maternal reported offspring symptoms, in the hyperactivity domain only. However, this was not replicated in ALSPAC where mothers had more prevalent daily alcohol use (16% in ALSPAC and 0.45% in MoBa). We did not observe associations of offspring GRS for alcohol use and metabolism with high risk of maternal or teacher reported childhood ADHD symptoms, in any of the cohorts, or for any of the ADHD symptom domains, even after adjustment for maternal and paternal GRS where this was available.

Our findings are somewhat dissimilar from previous findings in ALSPAC which found the risk of early onset conduct problems and decreased IQ to be associated with the *ADH* variants, particularly in children whose mothers drank during pregnancy (Lewis et al., 2012; Murray et al., 2016). However, these studies used a limited number of offspring genetic variants in *ADH* genes as a proxy for fetal alcohol exposure. The present study was based on larger numbers of mother-offspring dyads (4x more), and used all available *ADH/ALDH* variants. Other studies that have examined the association between maternal PAE and behavioural problems in offspring using quasi-experimental design to account for shared genetic and environmental factors found evidence for a potential causal effect on conduct disorder symptoms in childhood, and emotional reactiveness at age 3 (reduced aggressiveness at age 5) but not with ADHD symptoms in any of the studies (D’Onofrio et al., 2007; Lund et al., 2019). However, another study using a similar design in the MoBa cohort observed an association with ADHD symptoms at age 5 years measured with CPRS-R, but not with CBCL or when ADHD diagnosis was used (Eilertsen et al., 2017). Although in the studies that used a sibling comparison design, the association may be still affected by non-shared environmental confounders. The genetic variants used in our study can help to overcome the limitation of unmeasured confounding. Triangulating findings across the studies based on different study designs which have a different source of bias can provide more support whether a causal relationship exists. Given that the results are not consistent across the studies, no strong conclusions can be made.

In addition, a common limitation of previous studies reporting a positive association between PAE, conduct disorder and ADHD symptoms (eg Murray et al. (2016), Lund et al. (2019), D’Onofrio et al. (2007) and Eilertsen et al., 2017) is that they were based on outcomes that were measured only using maternal report. We have previously found inconsistent results for a potential causal effect when offspring ADHD symptoms were reported by mothers compared to teachers (Haan et al., 2021). This is not unexpected, as several studies had previously shown informant discrepancies in the assessment of children mental health problems which may be affected by factors such as mothers own mental health and socioeconomic status (Collishaw et al., 2009; De Los Reyes and Kazdin, 2005). Different findings in previous studies and our study may be also related to scales used for ADHD symptom assessment as observed in our previous study (Haan et al., 2021) and the study by Eilertsen and colleagues. The child’s age at the time of assessment and severity of the ADHD symptoms may as well have an impact on the observed results. It has been shown that presentation and manifestation of ADHD and other behavioural symptoms and diagnosis change from preschool to adolescence (Bunte et al., 2014; Curchack-Lichtin et al., 2014; Martel et al., 2017). This may lead to different findings across development as ADHD symptoms have been found to decline with age (Faraone et al., 2006). It is therefore possible that observed associations in previous studies may be influenced by reporter bias, child age and scales used for ADHD assessment and methods that did not sufficiently account for unmeasured confounds.

The major strength of this study is using a more comprehensive approach for identifying genetic variants affecting alcohol metabolism. We also compared findings across three international longitudinal cohorts of which one cohort includes genetic data from both parents and offspring. Having trio genetic data enables us to properly account for shared genetics and overcome potential biases such as dynastic effects and assortative mating (Davies et al., 2019).

However, several limitations should also be considered. First, our sample size may have been too small to detect a true effect of fetal alcohol exposure on offspring ADHD risk. MR studies require large sample sizes that can be difficult to gain in the context of intergenerational research and particularly in pregnancy substance use exposures, as many women reduce/stop using substances when planning pregnancy and while being pregnant. Second, although MR and GRS analyses are less affected by confounding, there are 3 assumptions that need to be met: 1) genetic variants are associated with the exposure; 2) genetic variants are not associated with the confounders; 3) genetic variants are not independently associated with the outcome. Considering that it is not possible to directly measure the level of alcohol the fetus is exposed to, we could not test the assumption 1 directly, as well as and assumptions 2 and 3 cannot be formally verified. Therefore, the risk of horizontal pleiotropy (violation of assumption 3) where the genetic variant directly affects the outcome independently of the exposure of interest still remains. Third, it is not possible to measure fetal blood alcohol levels to examine whether a dose-response relationship exists. Fourth, maternal PAE was based on self-reports and mothers may have underreported their prenatal alcohol use due to social desirability bias. This may have caused bias in the offspring GRS analyses where we stratified analyses based on maternal drinking status during pregnancy. Fifth, it is possible that lack of effects could be also influenced by dichotomizing outcome measures. Very often, dichotomized measures are used as the scales used in ADHD assessment (SDQ, CBCL, CPRS-R) generate skewed distributions (Swanson et al., 2012). Considering that research suggests that ADHD symptoms across a population are continuously distributed (Ahmad and Hinshaw, 2017; Larsson et al., 2012), continuous measures associated with ADHD symptoms could be more sensitive measures to use for investigating effects of fetal alcohol exposure. Sixth, longitudinal birth cohorts may be affected by selection bias if some groups are under-represented in the initial recruitment. Attrition over time can also affect representativeness which may cause biased exposure-outcome associations (Munafò et al., 2018). For example, in MoBa women who smoked during pregnancy, lived alone and had two previous births were under-represented (Nilsen et al., 2009) and in GenR women who continued to participate in the study were older and better educated (Kooijman et al., 2016). In addition, analyses restricted to live births such as the present one may incur in some degree of selection bias, in the form of collider bias (Liew et al., 2015), although it has been found that live birth bias is unlikely to affect studies investigating perinatal factors (Heinke et al., 2020).

## Conclusions

We did not find evidence for a causal effect of fetal alcohol exposure on offspring ADHD risk in the general population. Considering that previous studies have found that PAE can affect a wide range of neurodevelopmental domains (Mattson et al., 2019), future studies should use a multidimensional approach and measure multiple neurodevelopmental outcomes at the same time. It has been also suggested that combining genetic and epigenetic data would allow to better distinguish FASD profile (Lange et al., 2019). In addition, larger samples with trio genetic data would increase power to detect a potential causal effect of maternal environmental exposures (including fetal alcohol exposure) on offspring outcomes.

## Data Availability

The informed consent obtained from ALSPAC participants does not allow the data to be made freely available through any third party maintained public repository. However, data used for this submission can be made available on request to the ALSPAC Executive. The ALSPAC data management plan describes in detail the policy regarding data sharing, which is through a system of managed open access.
The GenR data that support the findings of this study are available from the data management team of Generation R, but restrictions apply to the availability of these data, which were used under license for the current study and are therefore not publicly available. Data are, however, available from the authors upon reasonable request and with permission from the Generation R Management Team. Interested researchers can contact
In MoBa, the consent given by the participants does not open for storage of data on an individual level in repositories or journals. Researchers who want access to data sets for replication should submit an application to. Access to data sets requires approval from The Regional Committee for Medical and Health Research Ethics in Norway and an agreement with MoBa.

http://www.bristol.ac.uk/alspac/researchers/access/

http://www.bristol.ac.uk/alspac/researchers/our-data/

## References

Achenbach, T. M., & Rescorla, L. A. (2001). Manual for the ASEBA School-Age Forms & Profiles. Burlington: University of Vermont, Research Centre for Children, Youth and Families.

Ahmad, S., & Hinshaw, S. (2017). Attention-Deficit/Hyperactivity Disorder, Trait Impulsivity, and Externalizing Behavior in a Longitudinal Sample. Journal of Abnormal Child Psychology, 45(6), 1077-1089. doi:http://dx.doi.org/10.1007/s10802-016-0226-9

Arnold, M., Raffler, J., Pfeufer, A., Suhre, K., & Kastenmuller, G. (2015). SNiPA: an interactive, genetic variant-centered annotation browser. Bioinformatics, 31(8), 1334–1336. doi:10.1093/bioinformatics/btu779

Aylward, G. P., & Stancin, T. (2008). Screening and assessment tools. Measurement and psychometric considerations. In Developmental Behavioral Pediatrics.Evidence and Practice (pp. 123–129). Philadelphia: Elsiver.

Birley, A. J., James, M. R., Dickson, P. A., Montgomery, G. W., Heath, A. C., Martin, N. G., & Whitfield, J. B. (2009). ADH single nucleotide polymorphism associations with alcohol metabolism in vivo. Human Molecular Genetics, 18(8), 1533–1542.

Birley, A. J., James, M. R., Dickson, P. A., Montgomery, G. W., Heath, A. C., Whitfield, J. B., & Martin, N. G. (2008). Association of the gastric alcohol dehydrogenase gene ADH7 with variation in alcohol metabolism. Human Molecular Genetics, 17(2), 179–189. doi:10.1093/hmg/ddm295

Boyd, A., Golding, J., Macleod, J., et al. (2013). Cohort Profile: the ‘children of the 90s’--the index offspring of the Avon Longitudinal Study of Parents and Children. International Journal of Epidemiology, 42(1), 111–127. doi:10.1093/ije/dys064

Bunte, T. L., Schoemaker, K., Hessen, D. J., van der Heijden, P. G. M., & Matthys, W. (2014). Stability and Change of ODD, CD and ADHD Diagnosis in Referred Preschool Children. Journal of Abnormal Child Psychology, 42(7), 1213–1224. doi:10.1007/s10802-014-9869-6

Burd, L. (2016). FASD and ADHD: Are they related and How? Bmc Psychiatry, 16. doi:ARTN 325 10.1186/s12888-016-1028-x

Burd, L., Blair, J., & Dropps, K. (2012). Prenatal alcohol exposure, blood alcohol concentrations and alcohol elimination rates for the mother, fetus and newborn. Journal of Perinatology, 32(9), 652–659. doi:10.1038/jp.2012.57

Collishaw, S., Goodman, R., Ford, T., Rabe-Hesketh, S., & Pickles, A. (2009). How far are associations between child, family and community factors and child psychopathology informant-specific and informant-general? Journal of Child Psychology and Psychiatry, 50(5), 571–580. doi:10.1111/j.1469-7610.2008.02026.x

Curchack-Lichtin, J. T., Chacko, A., & Halperin, J. M. (2014). Changes in ADHD Symptom Endorsement: Preschool to School Age. Journal of Abnormal Child Psychology, 42(6), 993–1004. doi:10.1007/s10802-013-9834-9

D’Onofrio, B. M., Van Hulle, C. A., Waldman, I. D., Rodgers, J. L., Rathouz, P. J., & Lahey, B. B. (2007). Causal inferences regarding prenatal alcohol exposure and childhood externalizing problems. Archives of General Psychiatry, 64(11), 1296–1304. doi:DOI 10.1001/archpsyc.64.11.1296

Davey Smith, G., & Ebrahim, S. (2003). ‘Mendelian randomization’: can genetic epidemiology contribute to understanding environmental determinants of disease? International Journal of Epidemiology, 32(1), 1–22. doi:10.1093/ije/dyg070

Davey Smith, G., & Hemani, G. (2014). Mendelian randomization: Genetic anchors for causal inference in epidemiological studies. Human Molecular Genetics, 23(R1), R89-R98. doi:http://dx.doi.org/10.1093/hmg/ddu328

Davies, N., Howe, L., Brumpton, B., Havdahl, A., Evans, D. M., & Davey Smith, G. (2019). Within family Mendelian randomization studies. Human Molecular Genetics, 28(R2), R170-R179. doi:http://dx.doi.org/10.1093/hmg/ddz204

De Los Reyes, A., & Kazdin, A. E. (2005). Informant discrepancies in the assessment of childhood psychopathology: A critical review, theoretical framework, and recommendations for further study. Psychological Bulletin, 131(4), 483–509. doi:10.1037/0033-2909.131.4.483

Easey, K. E., Dyer, M. L., Timpson, N. J., & Munafò, M. R. (2019). Prenatal alcohol exposure and offspring mental health: A systematic review. Drug Alcohol Depend. doi:10.1016/j.drugalcdep.2019.01.007

Edenberg, H. J., & McClintick, J. N. (2018). Alcohol Dehydrogenases, Aldehyde Dehydrogenases, and Alcohol Use Disorders: A Critical Review. Alcoholism-Clinical and Experimental Research, 42(12), 2281–2297. doi:10.1111/acer.13904

Eilertsen, E. M., Gjerde, L. C., Reichborn-Kjennerud, T., et al. (2017). Maternal alcohol use during pregnancy and offspring attention-deficit hyperactivity disorder (ADHD): a prospective sibling control study. International Journal of Epidemiology, 46(5), 1633–1640. doi:10.1093/ije/dyx067

Faraone, S. V., Biederman, J., & Mick, E. (2006). The age-dependent decline of attention deficit hyperactivity disorder: a meta-analysis of follow-up studies. Psychological Medicine, 36(2), 159–165. doi:10.1017/S003329170500471x

Fraser, A., Macdonald-Wallis, C., Tilling, K., et al. (2013). Cohort Profile: The Avon Longitudinal Study of Parents and Children: ALSPAC mothers cohort. International Journal of Epidemiology, 42(1), 97–110. doi:10.1093/ije/dys066

Haan, E., Sallis, H., Swanson, J., Ystrom, E., Munafò, M., Havdahl, A., & Zuccolo, L. (2019). Mendelian Randomization of prenatal alcohol exposure and ADHD in offspring. Open Science Framework. doi:https://doi.org/10.17605/OSF.IO/AQRXP

Haan, E., Sallis, H. M., Zuccolo, L., Labrecque, J., Ystrom, E., Reichborn-Kjennerud, T., Andreassen, O., Havdahl, A., & Munafò, M. (2021). Prenatal smoking, alcohol and caffeine exposure and ADHD risk in childhood: parental comparisons and polygenic risk score (PRS) analyses. medRxiv. doi:https://doi.org/10.1101/2021.03.25.21254087

Heinke, D., Rich-Edwards, J. W., Williams, P. L., et al. (2020). Quantification of selection bias in studies of risk factors for birth defects among livebirths. Paediatric and Perinatal Epidemiology, 34(6), 655–664. doi:10.1111/ppe.12650

Kapushesky, M., Emam, I., Holloway, E., et al. (2010). Gene Expression Atlas at the European Bioinformatics Institute. Nucleic Acids Research, 38, D690–D698. doi:10.1093/nar/gkp936

Kelly, Y., Iacovou, M., Quigley, M., Gray, R., Wolke, D., Kelly, J., & Sacker, A. (2013). Light drinking versus abstinence in pregnancy - behavioural and cognitive outcomes in 7-year-old children: a longitudinal cohort study. Bjog-an International Journal of Obstetrics and Gynaecology, 120(11), 1340–1347. doi:10.1111/1471-0528.12246

Kooijman, M. N., Kruithof, C. J., van Duijn, C. M., et al. (2016). The Generation R Study: design and cohort update 2017. European Journal of Epidemiology, 31(12), 1243–1264. doi:10.1007/s10654-016-0224-9

Lange, S., Shield, K., Rehm, J., Anagnostou, E., & Popova, S. (2019). Fetal alcohol spectrum disorder: neurodevelopmentally and behaviorally indistinguishable from other neurodevelopmental disorders. Bmc Psychiatry, 19(1). doi:ARTN 322 10.1186/s12888-019-2289-y

Larsson, H., Anckarsater, H., Rastam, M., Chang, Z., & Lichtenstein, P. (2012). Childhood attention-deficit hyperactivity disorder as an extreme of a continuous trait: a quantitative genetic study of 8,500 twin pairs. Journal of child psychology and psychiatry, and allied disciplines, 53(1), 73–80.

Lewis, S. J., Zuccolo, L., Davey Smith, G., et al. (2012). Fetal Alcohol Exposure and IQ at Age 8: Evidence from a Population-Based Birth-Cohort Study. Plos One, 7(11). doi:ARTN e49407 10.1371/journal.pone.0049407

Liew, Z., Olsen, J., Cui, X., Ritz, B., & Arah, O. A. (2015). Bias from conditioning on live-births in pregnancy cohorts: an illustration based on neurodevelopment in children after prenatal exposure to organic pollutants Response. International Journal of Epidemiology, 44(3), 1080–1081. doi:10.1093/ije/dyv140

Liu, M. Z., Jiang, Y., Wedow, R., et al. (2019). Association studies of up to 1.2 million individuals yield new insights into the genetic etiology of tobacco and alcohol use. Nature Genetics, 51(2), 237-+. doi:10.1038/s41588-018-0307-5

Lund, I. O., Eilertsen, E. M., Gjerde, L. C., Roysamb, E., Wood, M., Reichborn-Kjennerud, T., & Ystrom, E. (2019). Is the association between maternal alcohol consumption in pregnancy and pre-school child behavioural and emotional problems causal? Multiple approaches for controlling unmeasured confounding. Addiction, 114(6), 1004–1014. doi:10.1111/add.14573

Magnus, P., Birke, C., Vejrup, K., et al. (2016). Cohort Profile Update: The Norwegian Mother and Child Cohort Study (MoBa). International Journal of Epidemiology, 45(2), 382–388. doi:10.1093/ije/dyw029

Mamluk, L., Edwards, H. B., Savovic, J., et al. (2017). Low alcohol consumption and pregnancy and childhood outcomes: time to change guidelines indicating apparently ‘safe’ levels of alcohol during pregnancy? A systematic review and meta-analyses. Bmj Open, 7(7). doi:ARTN e015410 10.1136/bmjopen-2016-015410

Martel, M. M., Levinson, C. A., Lee, C. A., & Smith, T. E. (2017). Impulsivity Symptoms as Core to the Developmental Externalizing Spectrum. Journal of Abnormal Child Psychology, 45(1), 83–90. doi:10.1007/s10802-016-0148-6

Mattson, S. N., Bernes, G. A., & Doyle, L. R. (2019). Fetal Alcohol Spectrum Disorders: A Review of the Neurobehavioral Deficits Associated With Prenatal Alcohol Exposure. Alcoholism-Clinical and Experimental Research, 43(6), 1046–1062. doi:10.1111/acer.14040

Munafò, M., Tilling, K., Taylor, A. E., Evans, D. M., & Davey Smith, G. (2018). Collider scope: When selection bias can substantially influence observed associations. International Journal of Epidemiology, 47(1), 226-235. doi:http://dx.doi.org/10.1093/ije/dyx206

Murray, J., Burgess, S., Zuccolo, L., Hickman, M., Gray, R., & Lewis, S. J. (2016). Moderate alcohol drinking in pregnancy increases risk for children’s persistent conduct problems: causal effects in a Mendelian randomisation study. Journal of Child Psychology and Psychiatry, 57(5), 575–584. doi:10.1111/jcpp.12486

Niclasen, J., Andersen, A. M. N., Teasdale, T. W., & Strandberg-Larsen, K. (2014). Prenatal exposure to alcohol, and gender differences on child mental health at age seven years. Journal of Epidemiology and Community Health, 68(3), 224–232. doi:10.1136/jech-2013-202956

Nilsen, R., Vollset, S., Gjessing, H., et al. (2009). Self-selection and bias in a large prospective pregnancy cohort in Norway. Paediatric and Perinatal Epidemiology, 23(6), 597-608. doi:http://dx.doi.org/10.1111/j.1365-3016.2009.01062.x

Northstone, K., Lewcock, M., Groom, A., Boyd, A., Macleod, J., Timpson, N., & Wells, N. (2019). The Avon Longitudinal Study of Parents and Children (ALSPAC): an update on the enrolled sample of index children in 2019. Wellcome Open Res, 4, 51. doi:10.12688/wellcomeopenres.15132.1

Papatheodorou, I., Moreno, P., Manning, J., et al. (2020). Expression Atlas update: from tissues to single cells. Nucleic Acids Research, 48(D1), D77–D83. doi:10.1093/nar/gkz947

Patra, J., Bakker, R., Irving, H., Jaddoe, V. W. V., Malini, S., & Rehm, J. (2011). Dose-response relationship between alcohol consumption before and during pregnancy and the risks of low birthweight, preterm birth and small for gestational age (SGA)-a systematic review and meta-analyses. Bjog-an International Journal of Obstetrics and Gynaecology, 118(12), 1411–1421. doi:10.1111/j.1471-0528.2011.03050.x

Popova, S., Lange, S., Shield, K., Mihic, A., Chudley, A. E., Mukherjee, R. A. S., Bekmuradov, D., & Rehm, J. (2016). Comorbidity of fetal alcohol spectrum disorder: a systematic review and meta-analysis. Lancet, 387(10022), 978–987. doi:10.1016/S0140-6736(15)01345-8

Porter, M. S., Maravilla, J. C., Betts, K. S., & Alati, R. (2019). Low-moderate prenatal alcohol exposure and offspring attention-deficit hyperactivity disorder (ADHD): systematic review and meta-analysis. Archives of Gynecology and Obstetrics, 300(2), 269–277. doi:10.1007/s00404-019-05204-x

StataCorp. (2017). Stata Statistical Software: Release 15: College Station, TX: StataCorp LLC.

StataCorp. (2019). Stata Statistical Software: Release 16: College Station, TX: StataCorp LLC.

Swanson, J. M., Schuck, S., Porter, M. M., et al. (2012). Categorical and Dimensional Definitions and Evaluations of Symptoms of ADHD: History of the SNAP and the SWAN Rating Scales. Int J Educ Psychol Assess, 10(1), 51–70.

Weyrauch, D., Schwartz, M., Hart, B., Klug, M. G., & Burd, L. (2017). Comorbid Mental Disorders in Fetal Alcohol Spectrum Disorders: A Systematic Review. Journal of Developmental and Behavioral Pediatrics, 38(4), 283-291. doi:Doi 10.1097/Dbp.0000000000000440

