## Supplementary material for "“Maternal and offspring genetic risk score (GRS) analyses of fetal alcohol exposure and ADHD risk in offspring”"

### Study populations

In **ALSPAC**, the initial number of pregnancies enrolled is 14,541 and of these initial pregnancies, there was a total of 14,676 fetuses, resulting in 14,062 live births and 13,988 children who were alive at 1 year of age. When the oldest children were approximately 7 years of age, an attempt was made to bolster the initial sample with eligible cases who had failed to join the study originally, resulting in an additional 913 children being enrolled. The total sample size for analyses using any data collected after the age of seven is therefore 15,454 pregnancies, resulting in 15,589 fetuses. Of these 14,901 were alive at 1 year of age. The ALSPAC study was approved by the ALSPAC Ethics and Law Committee and the Local Research Ethics Committees and informed consent for the use of data collected via questionnaires and clinics was obtained from participants. The study website contains details of all the data that is available through a fully searchable data dictionary and variable search tool: http://www.bristol.ac.uk/alspac/researchers/our-data/

**GenR** is a population-based prospective cohort study in Rotterdam in the Netherlands designed to investigate environmental and genetic causes of health and development from fetal life until young adulthood. GenR recruited in total 9,778 pregnant women who were expected to give a birth between April 2002 and January 2006 in Rotterdam. Of all eligible children at birth, 61% agreed to participate in the study. The final baseline sample size included 9,749 children and 80% of the sample has been followed up until age 13 years (Kooijman et al., 2016). The GenR is a multi-ethnic cohort and besides Dutch ethnicity other larger ethnic groups are Surinamese, Turkish and Moroccan. The study was approved by the local Medical Ethical Committee (MEC 198.782/2001/31). Written informed consent was obtained from all participating women.

### MoBa is a population-based pregnancy cohort study conducted by the Norwegian Institute of Public Health. Participants were recruited from all over Norway from 1999-2008. The women consented to participation in 41% of the pregnancies. The cohort now includes 114.500 children, 95.200 mothers and 75.200 fathers. The current study is based on version 12 of the quality-assured data files released for research on January 2019. The establishment of MoBa and initial data collection was based on a license from the Norwegian Data Protection Agency and approval from The Regional Committees for Medical and Health Research Ethics. The MoBa cohort is now based on regulations related to the Norwegian Health Registry Act. The current study was approved by The Regional Committees for Medical and Health Research Ethics (2016/1702).

### Genome-Wide Data and Quality Control

**Avon Longitudinal Study of Parents and Children (ALSPAC)**

DNA samples were collected from 11,343 children and 10,015 mothers.

ALSPAC children were genotyped using the Illumina HumanHap550 quad chip genotyping platforms. Individuals were excluded on the basis of gender mismatches; minimal or excessive heterozygosity; disproportionate levels of individual missingness (>3%) and insufficient sample replication (IBD < 0.8). SNPs with a minor allele frequency of < 1%, a call rate of < 95% or evidence for violations of Hardy-Weinberg equilibrium (p < 10^-7^) were removed. Related subjects that passed all other quality control thresholds were retained during subsequent phasing and imputation. 9,115 children and 500,527 SNPs passed these quality control filters.

ALSPAC mothers were genotyped using the Illumina human660W-quad array at Centre National de Génotypage (CNG) and genotypes were called with Illumina GenomeStudio. SNPs were removed if they displayed more than 5% missingness or a Hardy-Weinberg equilibrium (p < 10^-6^). Additionally, SNPs with a minor allele frequency of less than 1% were removed. Samples were excluded if they displayed more than 5% missingness, had indeterminate X chromosome heterozygosity or extreme autosomal heterozygosity.

Related subjects that passed all other quality control thresholds were retained during subsequent phasing and imputation. 9,048 mothers and 526,688 SNPs passed these quality control filters.

Population stratification in mothers and children were compared with Hapmap II (release 22) European descent (CEU), Han Chinese, Japanese and Yoruba reference populations; all individuals with non-European ancestry were removed.

After combining genotype data in the mothers and the children, SNPs with genotype missingness above 1% were removed due to poor quality (11,396 SNPs removed) and a further 321 subjects were removed due to potential ID mismatches. This resulted in a dataset of 17,842 subjects. Imputation of the target data was performed using Impute V2.2.2 against the 1000 genomes reference panel (Phase 1, Version 3) (all polymorphic SNPs excluding singletons), using all 2186 reference haplotypes (including non-Europeans).

The final dataset included 8,237 children and 8,196 mothers.

More details about the genotyping and quality control procedure can be found (Paternoster et al., 2011; Taylor et al., 2018).

**Generation R (GenR)**

Genotype data were either collected from cord blood at birth (Illumina 610K Quad Chip) or via vena puncture (Illumina 660K Quad Chip) for 5,908 children (Generation R-1) and additional 320 samples were collected during a visit to the research centre at age 6 years (Generation R-2). Variants were filtered for minor allele frequency (MAF < 0.01), Hardy‐–Weinberg disequilibrium (p < 10^-7^) and missing rate (> 0.05). Individuals were additionally filtered on relatedness, sex mismatch and a total of 178 samples with genotyping rates lower than 97.5 % were excluded from the final projects (Generation R-1 and Generation R-2 sets). The combined dataset, merged using only SNPs common to both platforms (n = 5809), consisted of 549,511 SNPs. Imputation was based on two different reference panels: HapMap Project Phase II Release 22, build 36 phasing and 1000 Genomes Project (phase III release version), build 37 phasing.

Individuals from European descent were selected within 4 standard deviations on the first four genetic principal components of the HapMap Phase II Northwestern European (CEU) population. The final genome-wide data is available for 5,732 children from different ethnic backgrounds and for 2,661 children from the European ethnicity.

Currently maternal genotype data is not available yet and is going through quality control procedure.

More details about the genotyping procedure and quality control can be found elsewhere (Medina-Gomez et al., 2015).

**Norwegian Mother, Father and Child cohort (MoBa)**

Approximately 17,000 trios from the Norwegian Mother, Father and Child cohort were genotyped in three batches. The first batch, comprising 20,664 individuals and 542,585 SNPs was genotyped at the NTNU Genomics Core Facility (Trondheim, Oslo) using the Illumina HumanCoreExome (Illumina, San Diego, USA) genotyping array, version 12 1.1. The second batch, comprising 12,874 individuals and 547,644 SNPs was genotyped at the NTNU Genomics Core Facility (Trondheim, Oslo) using the Illumina HumanCoreExome (Illumina, San Diego, USA) genotyping array, version24 1.0. The third batch, comprising 17,949 individuals and 692,367 SNPs, was genotyped at ERASMUS MC (the Netherlands) using the Illumina Global Screening Array (Illumina, San Diego, USA) version 24 1. Individuals were excluded if they had a genotyping call rate below 95% or autosomal heterozygosity greater than four standard deviations from the sample mean. SNPs were excluded if they were ambiguous (A / T and C / G), had a genotyping call rate below 98%, minor allele frequency of less than 1%, or Hardy-Weinberg equilibrium P-value less than 1 × 10^-6^. Relatedness was assessed by flagging one individual from each pairwise comparison of identity-by-descent with a pi-hat greater than 0.1.

Population stratification was assessed, using the HapMap phase 3 release 3 as a reference by principal component analysis using EIGENSTRAT version 6.1.4. Visual inspection identified a homogenous population of European ethnicity and individuals of non-European ethnicity were removed. Duplicate samples were removed, and each genotyping batch was split into parents and offspring. Quality control was then conducted by genotyping array in parents and offspring’s separately.

The parents and offspring’s datasets were then merged into one dataset per genotyping batch; keeping only the SNPs that passed quality control in both datasets. All individuals passing the genotyping call rate and autosomal heterozygosity measures were included in the merged datasets. Therefore, the merged datasets included individuals previously excluded or flagged as a duplicate, ethnic outlier, having a sex discrepancy, or high level of relatedness. Concordance checks were then conducted on validated duplicates. Duplicate, tri-allelic and discordant (any discordance between the validated duplicates) SNPs were excluded. Individuals and SNPs with a genotyping call rate below 98% in the merged datasets were excluded. The duplicate sample that was removed before the start of the quality control was then excluded. Mendelian errors identified by the assessment of duos and trios were then recoded to missing. Insertions and deletions were also excluded.

After QC the Human Core Exome 12 batch comprised 20,231 individuals and 384,855 SNPs, the Human Core Exome 24 batch comprised 12,757 individuals and 396,189 SNPs, and the Global Screening Array batch comprised 17,742 individuals and 568,275 SNPs. Imputation was conducted separately for each genotyping batch by using the Haplotype reference consortium (HRC) release 1-1 as the genetic reference panel. Post imputation quality control was performed by removing individuals if they had a genotyping call rate less than 99% or were of non-European ethnicity. After quality control, a core homogeneous sample of European ethnicity (based on PCA of markers overlapping with available HapMap markers), unrelated (within generation, defined as accumulated identity-by-descent <0.015 and overall identity-by-descent PI_HAT <10%) individuals across all batches and arrays were available for use in analysis (N_children_ = 15,208; N_mothers_ = 14,804; N_fathers_ = 15,198).

More details about the genotyping and quality control procedure has been reported elsewhere (Helgeland et al., 2019).

**Supplementary Table S1. Assessment of alcohol consumption during pregnancy**

| **Cohort** | **Assessment of alcohol consumption** |
| --- | --- |
| **ALSPAC** | Mothers were asked about their average amount and frequency of alcohol consumption in the 18 and 32 weeks of gestation.  In the 18 weeks of gestation alcohol consumption was classified as never, <1 unit/week, >1 unit/week, 1–2 units/day, 3–9 units/day, or 10+ units/day.  32 weeks of gestation mothers were asked about their weekday and weekend alcohol consumption in units from which total alcohol consumption per week was calculated.  *1 drink/unit corresponds to 8 grams of pure alcohol (Alati et al., 2013) |
| **GenR** | Mothers were asked about their alcohol use and frequency in the early (<18 weeks), mid (18-25 weeks) and late (>25 weeks) pregnancy.  Maternal alcohol consumption was classified: <1 drink per week; 1-3 drinks per week; 4-6 drinks per week; 1 drink per day; 2-3 drinks per day; >3 drinks per day  *1 alcoholic drink corresponds to 12 grams of pure alcohol (Bakker et al., 2010) |
| **MoBa** | Mothers were asked about their frequency and amount of alcohol use during the current pregnancy in the questionnaires completed around 17 and 30 weeks of gestation and 6 months after child’s birth.  Continuous measure of weekly alcohol consumption in units was derived.  *1 alcoholic drink corresponds to 12.8 grams of pure alcohol (Knudsen et al., 2014) |

**Supplementary Table S2. ADHD assessment**

| **Cohort** | **Instrument** |
| --- | --- |
| **ALSPAC** | **Maternal and teacher report of The Development And Well-Being Assessment (DAWBA)** at age 7.5 years. The DAWBA was designed to generate psychiatric diagnoses based on questionnaires and interviews and brings together different sources of information for predicting psychiatric problems in children and adolescence (Goodman et al., 2000). It consists of 18 items that are measuring ADHD and separately inattentive and hyperactive-impulsive symptom domains. The items are assessed in the 3-point Likert scale with a degree of symptom: 0=no more than other; 1=a little more than others and 2=lot more than others. In ALSPAC a questionnaire was used.  **Maternal and teacher report of The Strengths and Difficulties Questionnaire (SDQ)** at age 7.5 years. The SDQ is a screening questionnaire for measuring children’s behaviour, emotions and relationships (Goodman, 1997). The hyperactivity scale consists of 5 items. The items are assessed in the 3-point Likert scale in the presence of symptoms: 0=not true; 1=somewhat true, and 2=certainly true |
| **GenR** | **Maternal report of the revised Conner’s Parent Rating Scale (CGRS-R)** at age 7.5 years. The CGRS-R was developed for screening and assessing child’s behavioural problems based on parental report and has been found to be a good instrument for distinguishing ADHD symptom domains (hyperactive-impulsive and inattention) (Conners et al., 1998). It consists of 9 items measuring hyperactivity-impulsivity symptoms and 12 items measuring cognitive problems (focusing on attention problems). The items are assessed in the 4-point Likert scale (0 for not at all true to 3 for very much true).  **Maternal report of The Child Behaviour Checklist (CBCL)** at age 6 years. The CBCL provides information on child’s behavioral problems and social competencies (Achenbach et al., 2001). Attention problems subscale consists of 5 items.  **Teacher Report Form of CBCL (CBCL-TRF)** at age 7 years. TRF consists of 26 items.  The items in CBCL and TRF are assessed in the 3-point Likert scale in the presence of symptoms: 0=not true, 1=somewhat true and 2= very true. |
| **MoBA** | **Maternal report of the Parent/Teacher Rating Scale for Disruptive Behaviour Disorders (RS-DBD)** at age 8. The RS-DBD has a parent and teacher version for measuring disruptive behaviour disorders in children. It consists of 18 items that are measuring ADHD symptoms and separately hyperactive-impulsive and inattentive symptom domains (Silva et al., 2005). The items are rated in the 4-point Likert scale: 1 = not at all; 2 = just a little; 3 = pretty much and 4 = very much.  In MoBa, currently maternal report is available. |
|  | All the included scales have shown good psychometric properties (Conners et al., 1998; Goodman et al., 2000; Silva et al., 2005), although in SDQ and CBCL scales, the validity is somewhat lower and the rate of false positives is higher in terms of psychiatric disorders (Goodman, 2001; Rishel et al., 2005).  However, SDQ and CBCL are broadband screening tools with only a few items on ADHD symptoms, whereas the DAWBA questionnaire, CGRS-R and RS-DBD are specifically designed to identify symptoms of ADHD. |

### Supplementary Figure S1. SNPs positioned in chromosome 4 before clumping

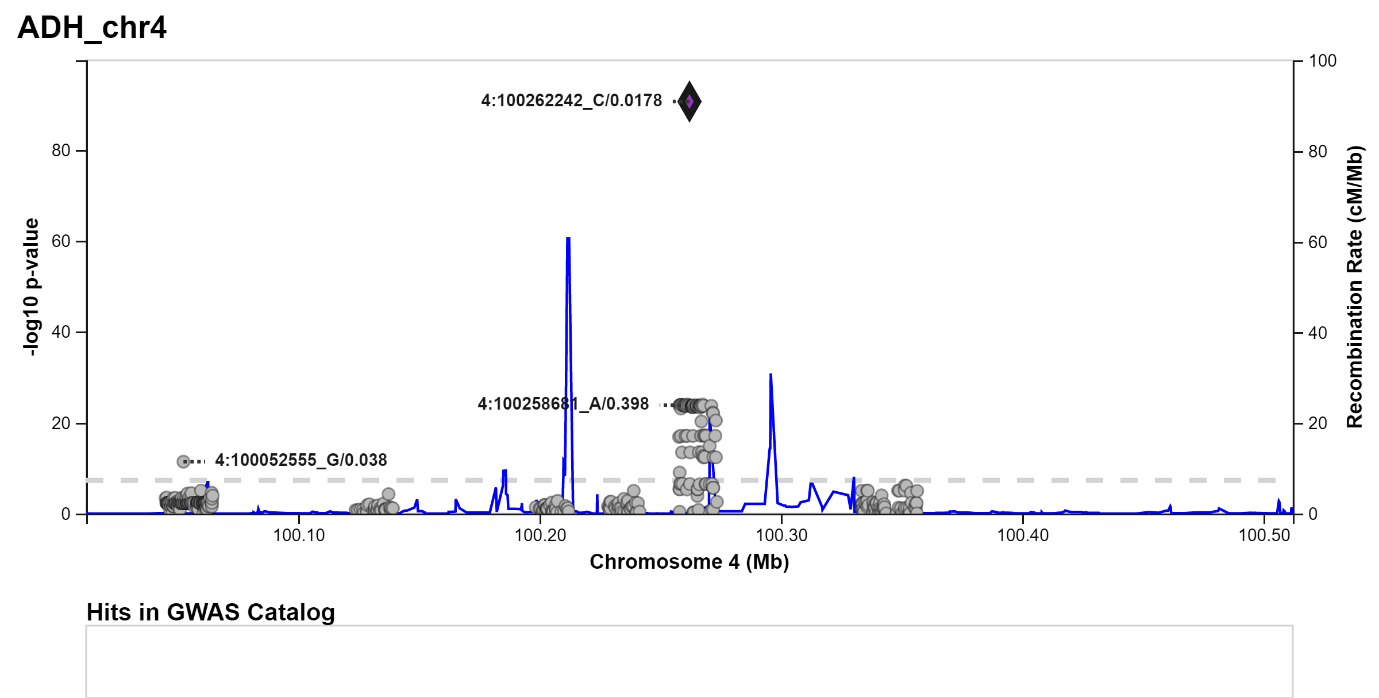

*Note: 551 SNPs were identified from ADH4, ADH5, ADH6, ADH7, ADH1A, ADH1B, ADH1C genes*

### Supplementary Figure S2. SNPs positioned in chromosome 9 before clumping

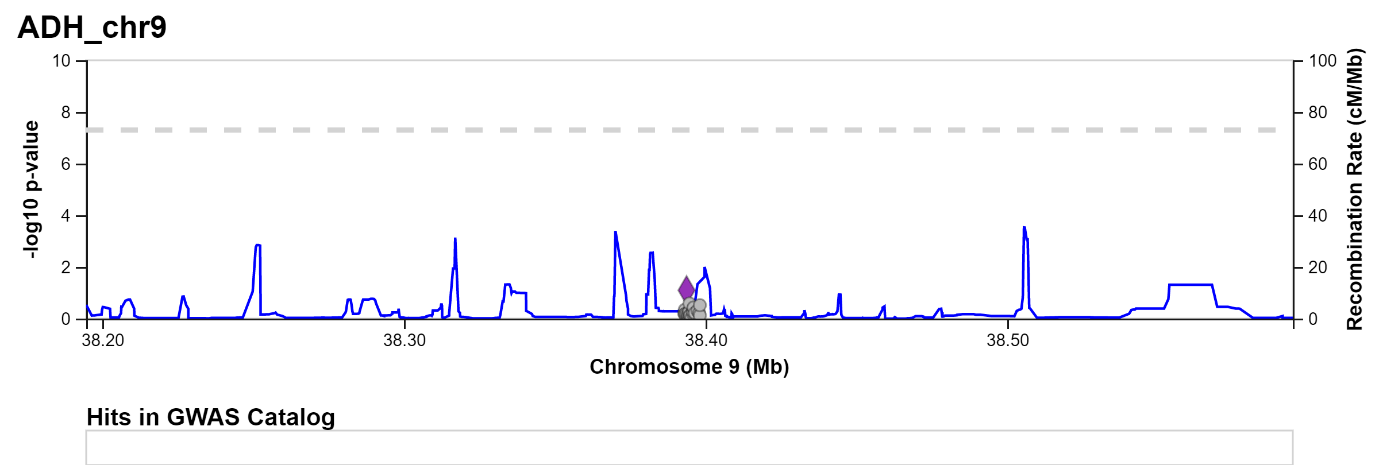

*Note: 280 SNPs were identified from ALDH1B1 and ALDH1A1 genes*

### Supplementary Figure S3. SNPs positioned in chromosome 12 before clumping

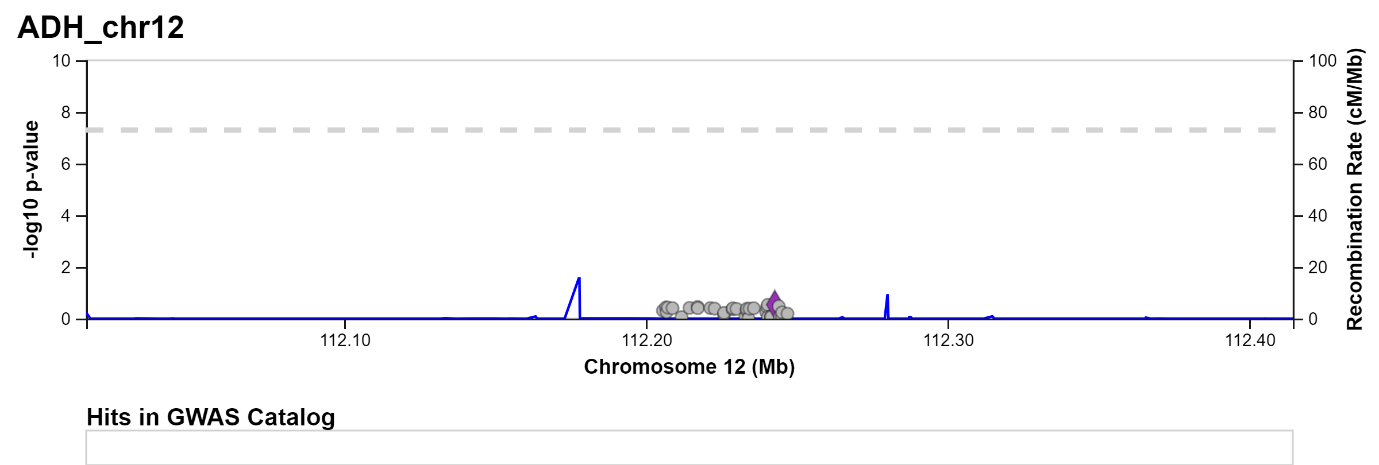

*Note: 38 SNPs were identified from ALDH2 gene*

**Supplementary Table S3. Independent SNPs identified after clumping**

| **SNP** | **Chromosome** |
| --- | --- |
| rs7669660 | 4 |
| rs116010022 | 4 |
| rs28730582 | 4 |
| rs29001207 | 4 |
| rs13125262 | 4 |
| rs17033 | 4 |
| rs138331988 | 4 |
| rs17028839 | 4 |
| rs138244919 | 4 |
| rs141973904 | 4 |
| rs3805329 | 4 |
| rs75756595 | 4 |
| rs1154465 | 4 |
| rs4646769 | 9 |
| rs77054814 | 9 |
| rs12378961 | 9 |
| rs10973779 | 9 |
| rs8187999 | 9 |
| rs8187996 | 9 |
| rs168351 | 9 |
| rs8187953 | 9 |
| rs8187950 | 9 |
| rs78094588 | 9 |
| rs8187928 | 9 |
| rs34878833 | 9 |
| rs8187898 | 9 |
| rs80105873 | 9 |
| rs8187891 | 9 |
| rs17648566 | 9 |
| rs116917518 | 9 |
| rs11143426 | 9 |
| rs41287405 | 9 |
| rs148620777 | 12 |
| rs2283354 | 12 |
| rs73205605 | 12 |
| rs61941278 | 12 |

**Supplementary Table S4. Harmonisation of SNPs in ALSPAC based on the GSCAN summary statistics**

|  | **GSCAN** | | | | **ALSPAC** | | | |
| --- | --- | --- | --- | --- | --- | --- | --- | --- |
| **SNP** | **Non-effect allele** | **Effect allele** | **Effect allele frequency** | **Beta** | **Minor allele** | **Major allele** | **Minor allele frequency** | **Effect allele after harmonisation** |
| rs7669660 | T | C | 0.136 | 0.0086 | C | T | 0.1481 | C |
| rs116010022 | C | A | 0.00942 | -0.0023 | A | C | 0.0131 | C |
| rs28730582 | C | T | 0.0318 | 0.0149 | T | C | 0.0294 | T |
| rs29001207 | G | C | 0.038 | -0.0334 | C | G | 0.0512 | G |
| rs13125262 | G | C | 0.0436 | 0.0160 | C | G | 0.0498 | C |
| rs17033 | T | C | 0.0881 | -0.0063 | C | T | 0.0880 | T |
| rs138331988 | G | A | 0.0192 | 0.0042 | A | G | 0.0180 | A |
| rs17028839 | A | G | 0.039 | 0.0031 | G | A | 0.0437 | G |
| rs138244919 | C | T | 0.0272 | 0.0052 | T | C | 0.0258 | T |
| rs141973904 | C | T | 0.0178 | -0.1990 | T | C | 0.0120 | C |
| rs3805329 | T | C | 0.0625 | 0.0105 | C | T | 0.0650 | C |
| rs75756595 | G | A | 0.0522 | 0.0015 | A | G | 0.0499 | A |
| rs1154465 | T | A | 0.0276 | 0.0061 | A | T | 0.0248 | A |
| rs4646769 | T | C | 0.857 | -0.0014 | T | C | 0.1372 | T |
| rs77054814 | A | G | 0.067 | 0.0017 | G | A | 0.0612 | G |
| rs12378961 | C | G | 0.0573 | 0.0068 | G | C | 0.0705 | G |
| rs10973779 | G | A | 0.0299 | -0.0067 | A | G | 0.0312 | G |
| rs8187999 | C | G | 0.0238 | 0.0111 | G | C | 0.0265 | G |
| rs8187996 | C | T | 0.0479 | -0.0030 | T | C | 0.0502 | C |
| rs168351 | A | G | 0.147 | -0.0039 | G | A | 0.1564 | A |
| rs8187953 | C | G | 0.0268 | 0.0011 | G | C | 0.0308 | G |
| rs8187950 | A | G | 0.0364 | -0.0033 | G | A | 0.0366 | A |
| rs78094588 | G | A | 0.0233 | 0.0031 | A | G | 0.0178 | A |
| rs8187928 | C | T | 0.0253 | 0.0103 | T | C | 0.0212 | T |
| rs34878833 | G | A | 0.0231 | 0.0139 | A | G | 0.0348 | A |
| rs8187898 | T | C | 0.0259 | 0.0038 | C | T | 0.0288 | C |
| rs80105873 | G | T | 0.0289 | -0.0029 | T | G | 0.0337 | G |
| rs8187891 | T | C | 0.0252 | 0.0002 | C | T | 0.0268 | C |
| rs17648566 | T | C | 0.0204 | -0.0093 | C | T | 0.0205 | T |
| rs116917518 | A | T | 0.0351 | -0.0020 | T | A | 0.0512 | A |
| rs11143426 | A | G | 0.0127 | 0.0147 | G | A | 0.0150 | G |
| rs41287405 | T | C | 0.0238 | 0.0057 | C | T | 0.0283 | C |
| rs148620777 | A | G | 0.0177 | -0.0048 | G | A | 0.0177 | A |
| rs2283354 | G | A | 0.174 | 0.0023 | A | G | 0.1742 | A |
| rs73205605 | G | A | 0.036 | -0.0018 | A | G | 0.0442 | G |
| rs61941278 | A | G | 0.013 | 0.0075 | G | A | 0.0260 | G |

*Note: GWAS & Sequencing Consortium of Alcohol and Nicotine use*

**Supplementary Table S5. Harmonisation of SNPs in GenR based on the GSCAN summary statistics**

|  | **GSCAN** | | | | **GenR** | | | |
| --- | --- | --- | --- | --- | --- | --- | --- | --- |
| **SNP** | **Non-effect allele** | **Effect allele** | **Effect allele frequency** | **Beta** | **Minor allele** | **Major allele** | **Major allele frequency** | **Effect allele after harmonisation** |
| rs7669660 | T | C | 0.136 | 0.0086 | C | T | 0.8480 | C |
| rs116010022 | C | A | 0.00942 | -0.0023 | A | C | 0.9921 | C |
| rs28730582 | C | T | 0.0318 | 0.0149 | T | C | 0.9707 | T |
| rs29001207 | G | C | 0.038 | -0.0334 | C | G | 0.9554 | G |
| rs13125262 | G | C | 0.0436 | 0.0160 | C | G | 0.9471 | C |
| rs17033 | T | C | 0.0881 | -0.0063 | C | T | 0.9137 | T |
| rs138331988 | G | A | 0.0192 | 0.0042 | A | G | 0.9826 | A |
| rs17028839 | A | G | 0.039 | 0.0031 | G | A | 0.9570 | G |
| rs138244919 | C | T | 0.0272 | 0.0052 | T | C | 0.9821 | T |
| rs141973904 | C | T | 0.0178 | -0.1990 | T | C | 0.9809 | C |
| rs3805329 | T | C | 0.0625 | 0.0105 | C | T | 0.9486 | C |
| rs75756595 | G | A | 0.0522 | 0.0015 | A | G | 0.9599 | A |
| rs1154465 | T | A | 0.0276 | 0.0061 | A | T | 0.9838 | A |
| rs4646769 | T | C | 0.857 | -0.0014 | T | C | 0.8085 | T |
| rs77054814 | A | G | 0.067 | 0.0017 | G | A | 0.9521 | G |
| rs12378961 | C | G | 0.0573 | 0.0068 | G | C | 0.9491 | G |
| rs10973779 | G | A | 0.0299 | -0.0067 | A | G | 0.9441 | G |
| rs8187999 | C | G | 0.0238 | 0.0111 | G | C | 0.9740 | G |
| rs8187996 | C | T | 0.0479 | -0.0030 | T | C | 0.9525 | C |
| rs168351 | A | G | 0.147 | -0.0039 | G | A | 0.8714 | A |
| rs8187953 | C | G | 0.0268 | 0.0011 | G | C | 0.9736 | G |
| rs8187950 | A | G | 0.0364 | -0.0033 | G | A | 0.9634 | A |
| rs78094588 | G | A | 0.0233 | 0.0031 | A | G | 0.9793 | A |
| rs8187928 | C | T | 0.0253 | 0.0103 | T | C | 0.9686 | T |
| rs34878833 | G | A | 0.0231 | 0.0139 | A | G | 0.9708 | A |
| rs8187898 | T | C | 0.0259 | 0.0038 | C | T | 0.9785 | C |
| rs80105873 | G | T | 0.0289 | -0.0029 | T | G | 0.9783 | G |
| rs8187891 | T | C | 0.0252 | 0.0002 | C | T | 0.9781 | C |
| rs17648566 | T | C | 0.0204 | -0.0093 | C | T | 0.9747 | T |
| rs116917518 | A | T | 0.0351 | -0.0020 | T | A | 0.9596 | A |
| rs11143426 | A | G | 0.0127 | 0.0147 | G | A | 0.9899 | G |
| rs41287405 | T | C | 0.0238 | 0.0057 | C | T | 0.9729 | C |
| rs148620777 | A | G | 0.0177 | -0.0048 | G | A | 0.9839 | A |
| rs2283354 | G | A | 0.174 | 0.0023 | A | G | 0.8045 | A |
| rs73205605 | G | A | 0.036 | -0.0018 | A | G | 0.9578 | G |
| rs61941278 | A | G | 0.013 | 0.0075 | G | A | 0.9856 | G |

*Note: GWAS & Sequencing Consortium of Alcohol and Nicotine use*

**Supplementary Table S6. Harmonisation of SNPs in MoBa based on the GSCAN summary statistics**

|  |  | **GSCAN** | | | | **MoBa** | | | |
| --- | --- | --- | --- | --- | --- | --- | --- | --- | --- |
| **SNP** | **New proxy SNP** | **Non-effect allele** | **Effect allele** | **Effect allele frequency** | **Beta** | **Reference allele** | **Alternate allele** | **Reference allele frequency** | **Effect allele after harmonisation** |
| rs7669660 |  | T | C | 0.136 | 0.0086 | T | C | 0.1364 | C |
| rs116010022 | rs11724783 | C | A | 0.425 | 0.0003 | C | A | 0.4252 | A |
| rs28730582 |  | C | T | 0.0318 | 0.0149 | C | T | 0.0318 | T |
| rs29001207 |  | G | C | 0.038 | -0.0334 | G | C | 0.0380 | G |
| rs13125262 | rs13133633 | C | G | 0.272 | 0.0061 | C | G | 0.2717 | G |
| rs17033 |  | T | C | 0.0881 | -0.0063 | T | C | 0.0881 | T |
| rs138331988 |  | G | A | 0.0192 | 0.0042 | G | A | 0.0192 | A |
| rs17028839 |  | A | G | 0.039 | 0.0031 | A | G | 0.0390 | G |
| rs138244919 |  | C | T | 0.0272 | 0.0052 | C | T | 0.0272 | T |
| rs141973904 | rs143502255 | C | T | 0.366 | -0.0023 | C | T | 0.3658 | C |
| rs3805329 |  | T | C | 0.0625 | 0.0105 | T | C | 0.0625 | C |
| rs75756595 |  | G | A | 0.0522 | 0.0015 | G | A | 0.0522 | A |
| rs1154465 |  | T | A | 0.0276 | 0.0061 | T | A | 0.0276 | A |
| rs4646769 |  | T | C | 0.857 | -0.0014 | T | C | 0.8574 | T |
| rs77054814 |  | A | G | 0.067 | 0.0017 | A | G | 0.0670 | G |
| rs12378961 |  | C | G | 0.0573 | 0.0068 | C | G | 0.0573 | G |
| rs10973779 |  | G | A | 0.0299 | -0.0067 | G | A | 0.0299 | G |
| rs8187999 |  | C | G | 0.0238 | 0.0111 | C | G | 0.0238 | G |
| rs8187996 |  | C | T | 0.0479 | -0.0030 | C | T | 0.0479 | C |
| rs168351 |  | A | G | 0.147 | -0.0039 | A | G | 0.1467 | A |
| rs8187953 | rs918836 | G | C | 0.312 | 0.0002 | G | C | 0.3116 | C |
| rs8187950 | rs8187924 | G | A | 0.501 | -0.0010 | G | A | 0.5008 | G |
| rs78094588 |  | G | A | 0.0233 | 0.0031 | G | A | 0.0233 | A |
| rs8187928 |  | C | T | 0.0253 | 0.0103 | C | T | 0.0253 | T |
| rs34878833 |  | G | A | 0.0231 | 0.0139 | G | A | 0.0231 | A |
| rs8187898 |  | T | C | 0.0259 | 0.0038 | T | C | 0.0259 | C |
| rs80105873 | rs7848927 | G | T | 0.559 | -0.0004 | G | T | 0.5588 | G |
| rs8187891 |  | T | C | 0.0252 | 0.0002 | T | C | 0.0252 | C |
| rs17648566 | rs7860944 | C | T | 0.604 | -0.0026 | C | T | 0.6040 | C |
| rs116917518 |  | A | T | 0.0351 | -0.0020 | A | T | 0.0351 | A |
| rs11143426 |  | A | G | 0.0127 | 0.0147 | A | G | 0.0127 | G |
| rs41287405 | rs4237253 | C | T | 0.5 | -0.0010 | C | T | 0.4995 | C |
| rs148620777 |  | A | G | 0.0177 | -0.0048 | A | G | 0.0177 | A |
| rs2283354 |  | G | A | 0.174 | 0.0023 | G | A | 0.1738 | A |
| rs73205605 |  | G | A | 0.036 | -0.0018 | G | A | 0.0360 | G |
| rs61941278 | rs61941274 | G | A | 0.0148 | 0.0073 | G | A | 0.0148 | A |

*Note: GWAS & Sequencing Consortium of Alcohol and Nicotine use*

**Supplementary Figure S4**

a) Collider bias in maternal GRS analyses

**Measured and unmeasured Confounding**

**Maternal genetic variants of alcohol use and metabolism**

**ADHD in offspring at age 7-8**

**Fetal alcohol exposure**

**Maternal alcohol use during pregnancy**

*Note: Conditioning on maternal drinking status during pregnancy induces a form of bias known as collider bias. In practice, this manifests as an artefactual association (red dashed arrow) between genetic variants of alcohol use and confounders.*

b) Collider bias in offspring GRS analyses

**Measured and unmeasured Confounding**

**ADHD in offspring at age 7-8**

**Fetal alcohol exposure**

**Maternal alcohol use during pregnancy**

**Offspring genetic variants of alcohol use and metabolism**

*Note: Conditioning on maternal drinking status during pregnancy in offspring GRS analyses does not induce collider bias as offspring genetic variants does not affect maternal alcohol use. However, adjustment for maternal GRS is still necessary given the shared genetics between child and mother.*

### Supplementary Figure S5. Leave-one-out analyses in ALSPAC with maternal GRS on high risk of maternal reported ADHD symptoms

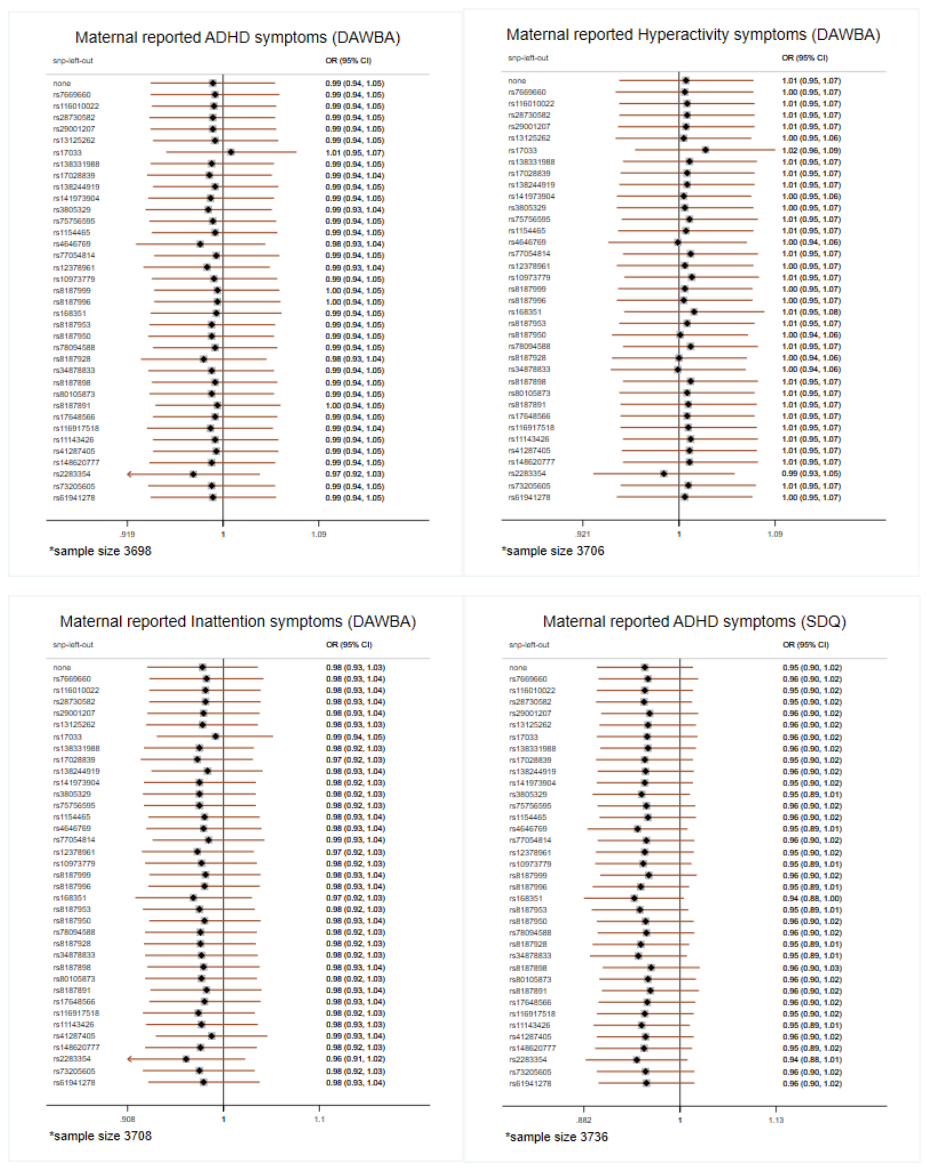

*Note: Development And Well-Being Assessment (DAWBA); Strength and Difficulties Questionnaire (SDQ)*

### Supplementary Figure S6. Leave-one-out analyses in ALSPAC with maternal GRS on high risk of teacher reported ADHD symptoms

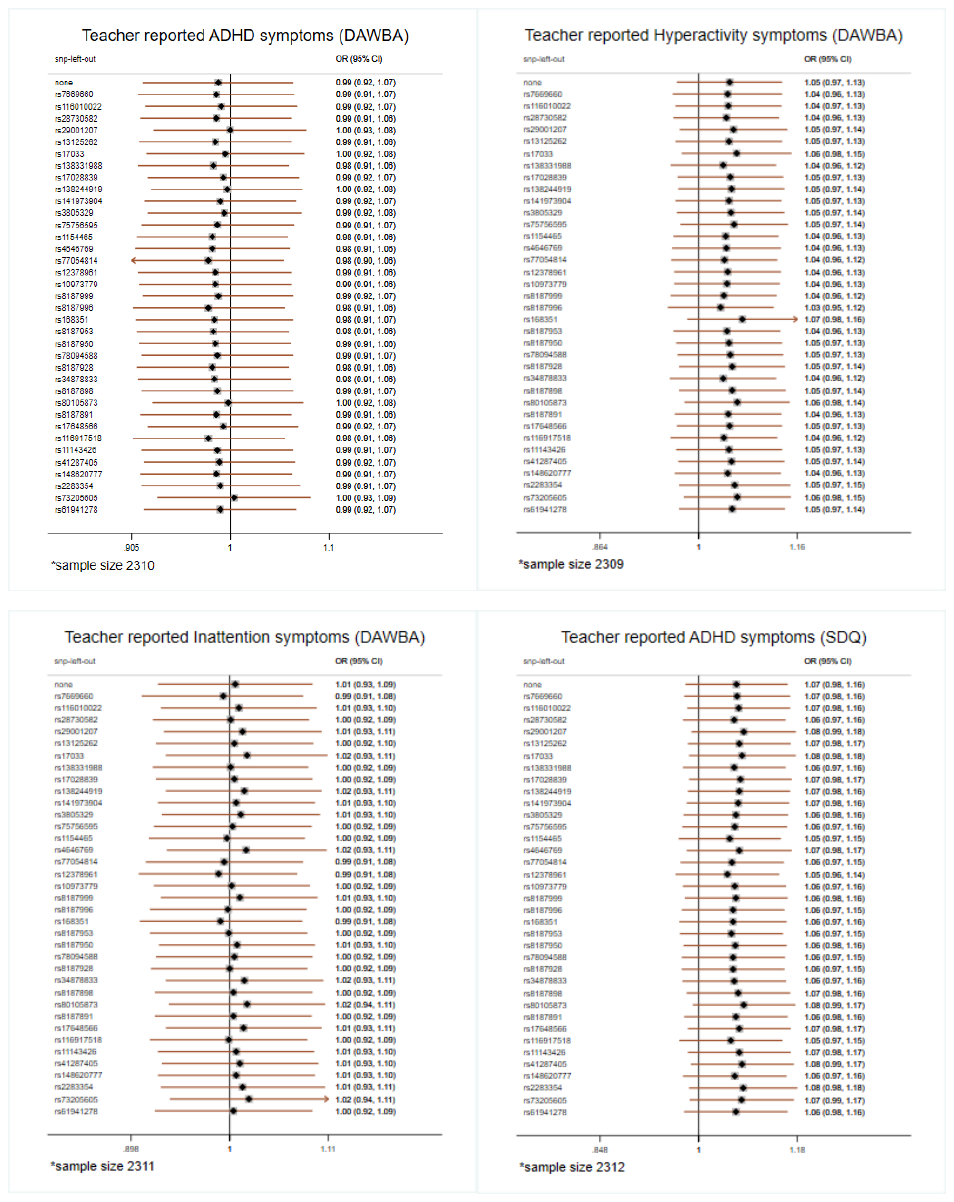

*Note: Development And Well-Being Assessment (DAWBA); Strength and Difficulties Questionnaire (SDQ)*

### Supplementary Figure S7. Leave-one-out analyses in ALSPAC with offspring GRS on high risk of maternal reported ADHD symptoms if mother drink during pregnancy

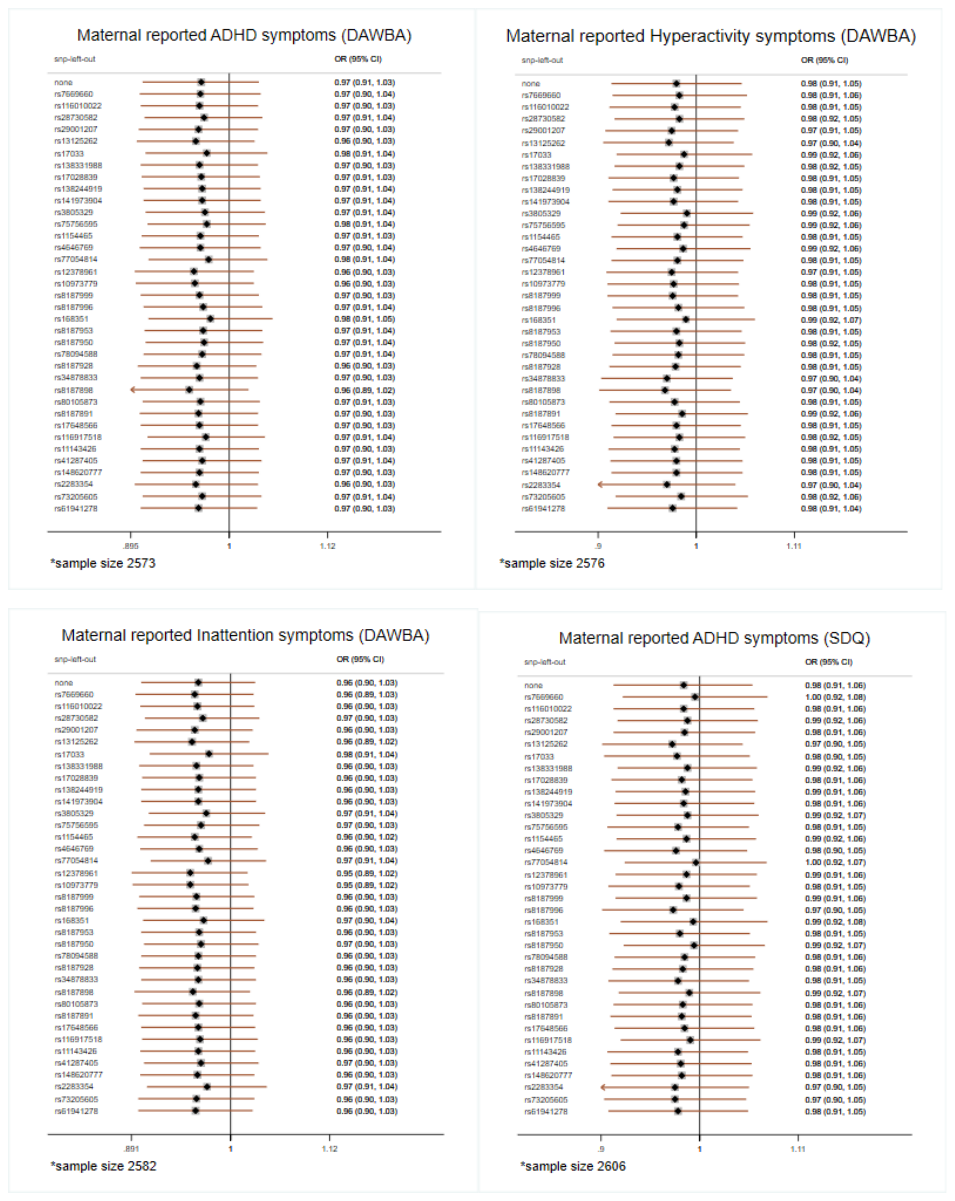

*Note: Development And Well-Being Assessment (DAWBA); Strength and Difficulties Questionnaire (SDQ)*

### Supplementary Figure S8. Leave-one-out analyses in ALSPAC with offspring GRS on high risk of maternal reported ADHD symptoms if mother did not drink during pregnancy

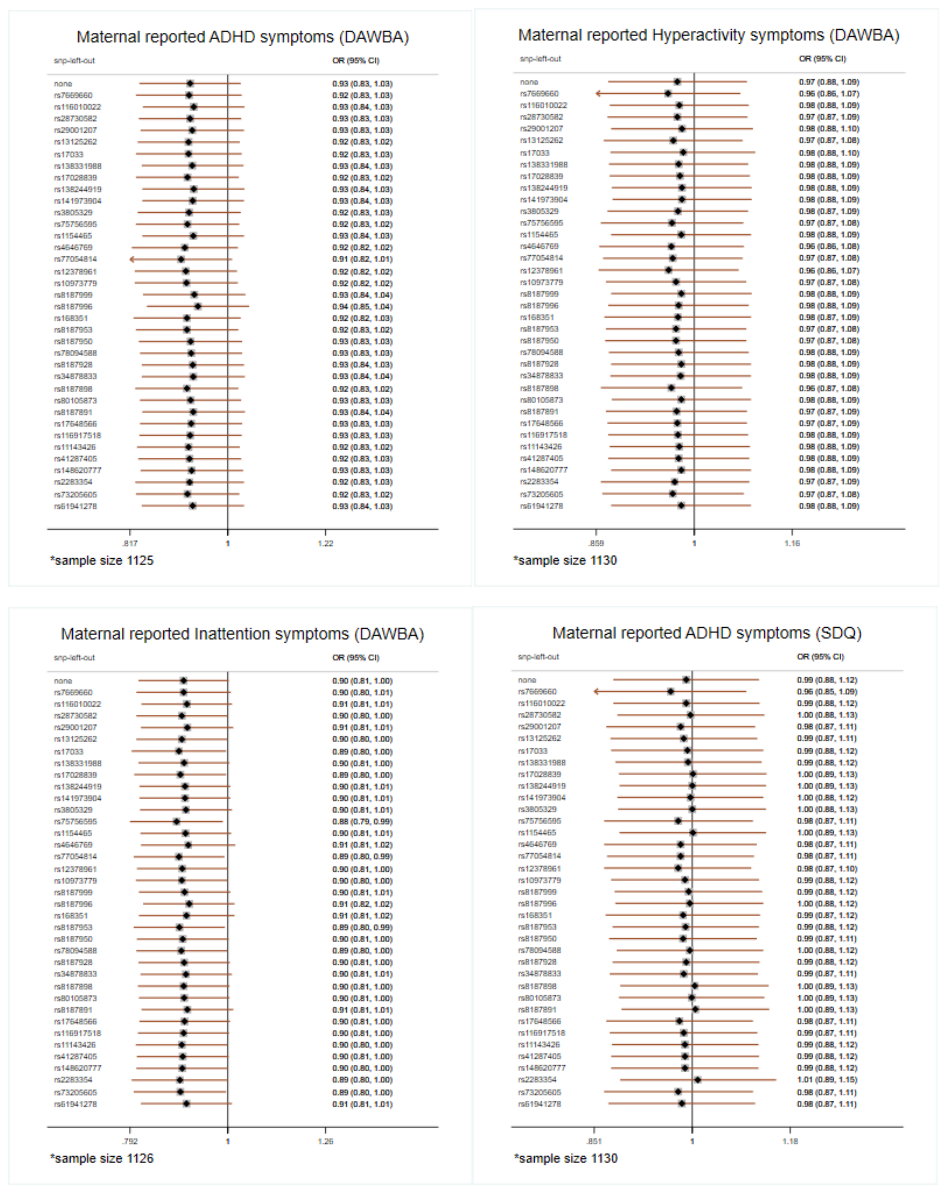

*Note: Development And Well-Being Assessment (DAWBA); Strength and Difficulties Questionnaire (SDQ)*

### Supplementary Figure S9. Leave-one-out analyses in ALSPAC with offspring GRS on high risk of teacher reported ADHD symptoms if mother drink during pregnancy

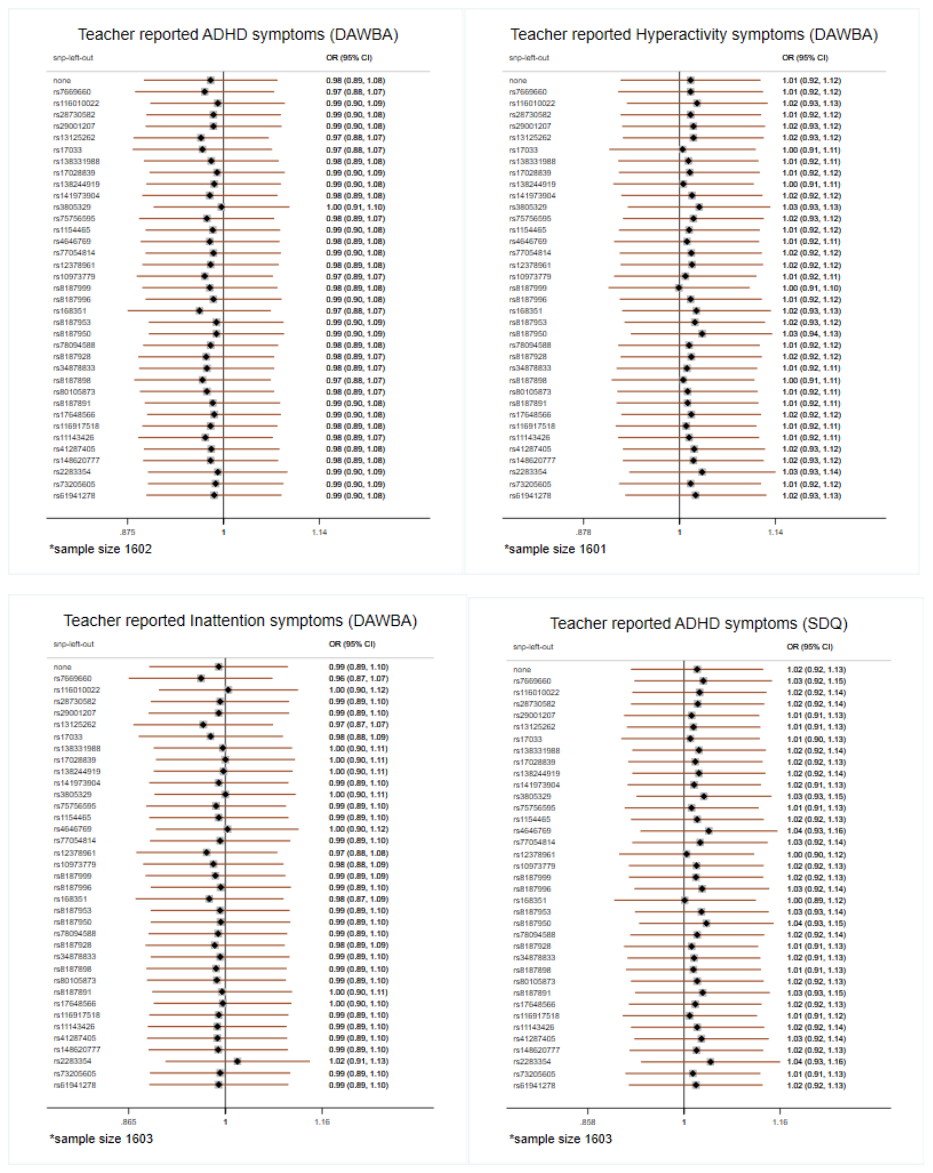

*Note: Development And Well-Being Assessment (DAWBA); Strength and Difficulties Questionnaire (SDQ)*

### Supplementary Figure S10. Leave-one-out analyses in ALSPAC with offspring GRS on high risk of teacher reported ADHD symptoms if mother did not drink during pregnancy

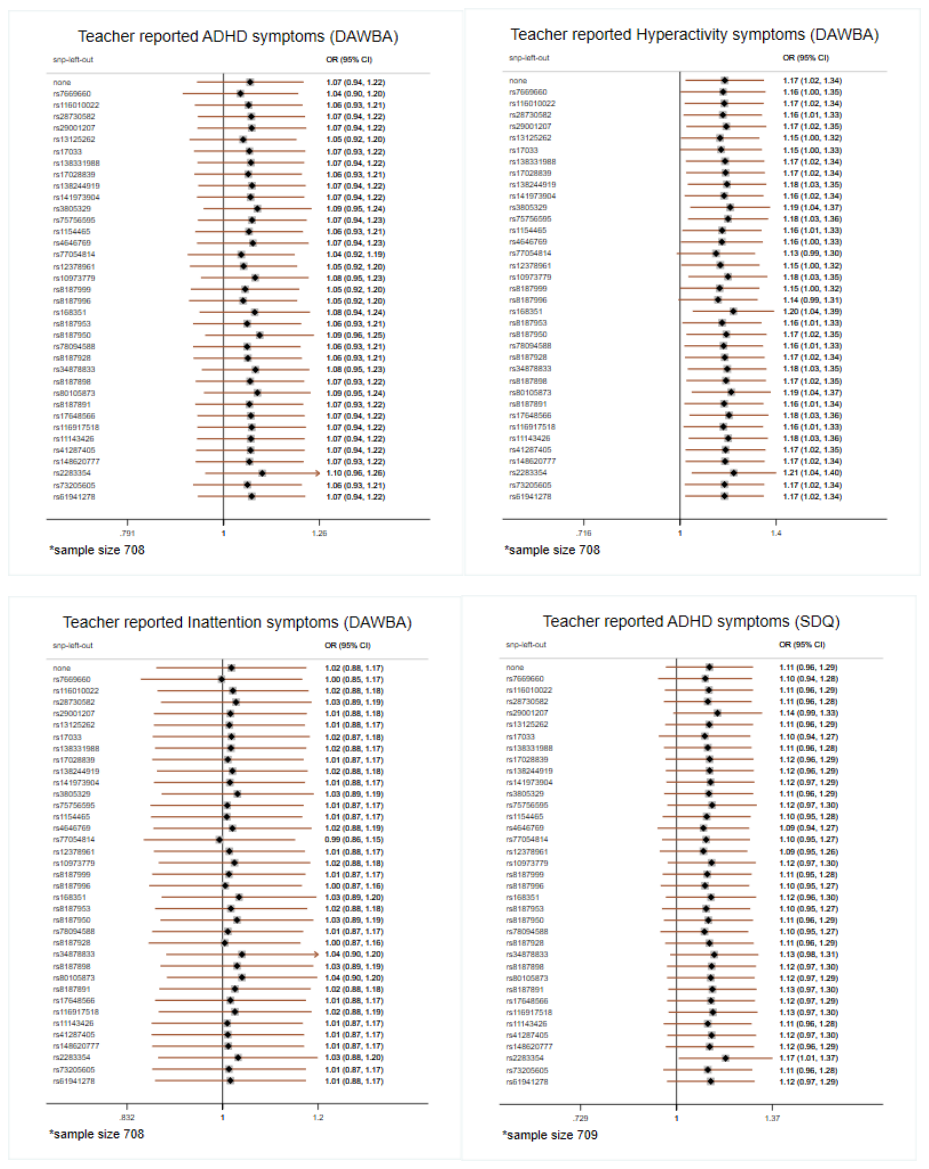

*Note: Development And Well-Being Assessment (DAWBA); Strength and Difficulties Questionnaire (SDQ)*

### Supplementary Figure S11. Leave-one-out analyses in GenR with offspring GRS on high risk of maternal and teacher reported ADHD symptoms if mother drink during pregnancy

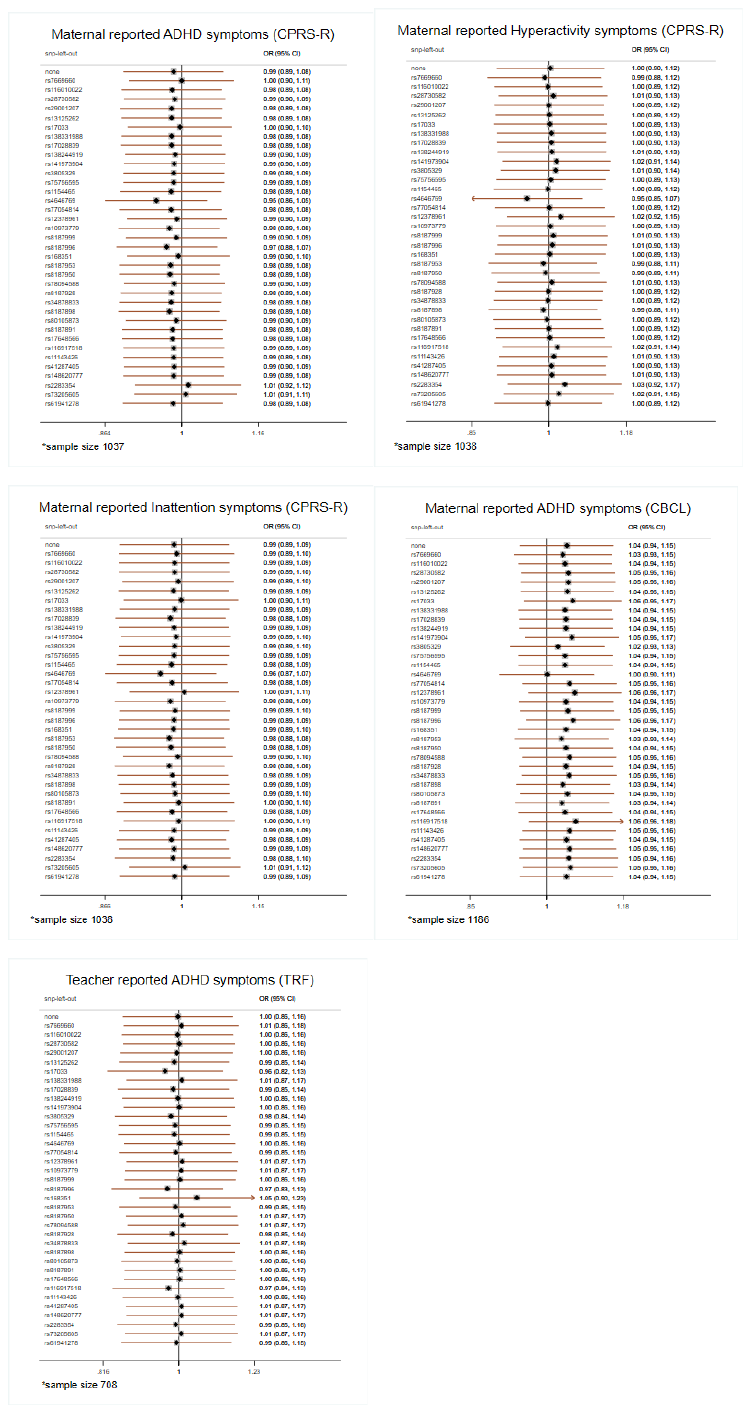

*Note: Revised Conner’s Parent Rating Scale (CGRS-R); Child Behaviour Checklist (CBCL); Teacher Report Form (TRF)*

### Supplementary Figure S12. Leave-one-out analyses in GenR with offspring GRS on high risk of maternal and teacher reported ADHD symptoms if mother did not drink during pregnancy

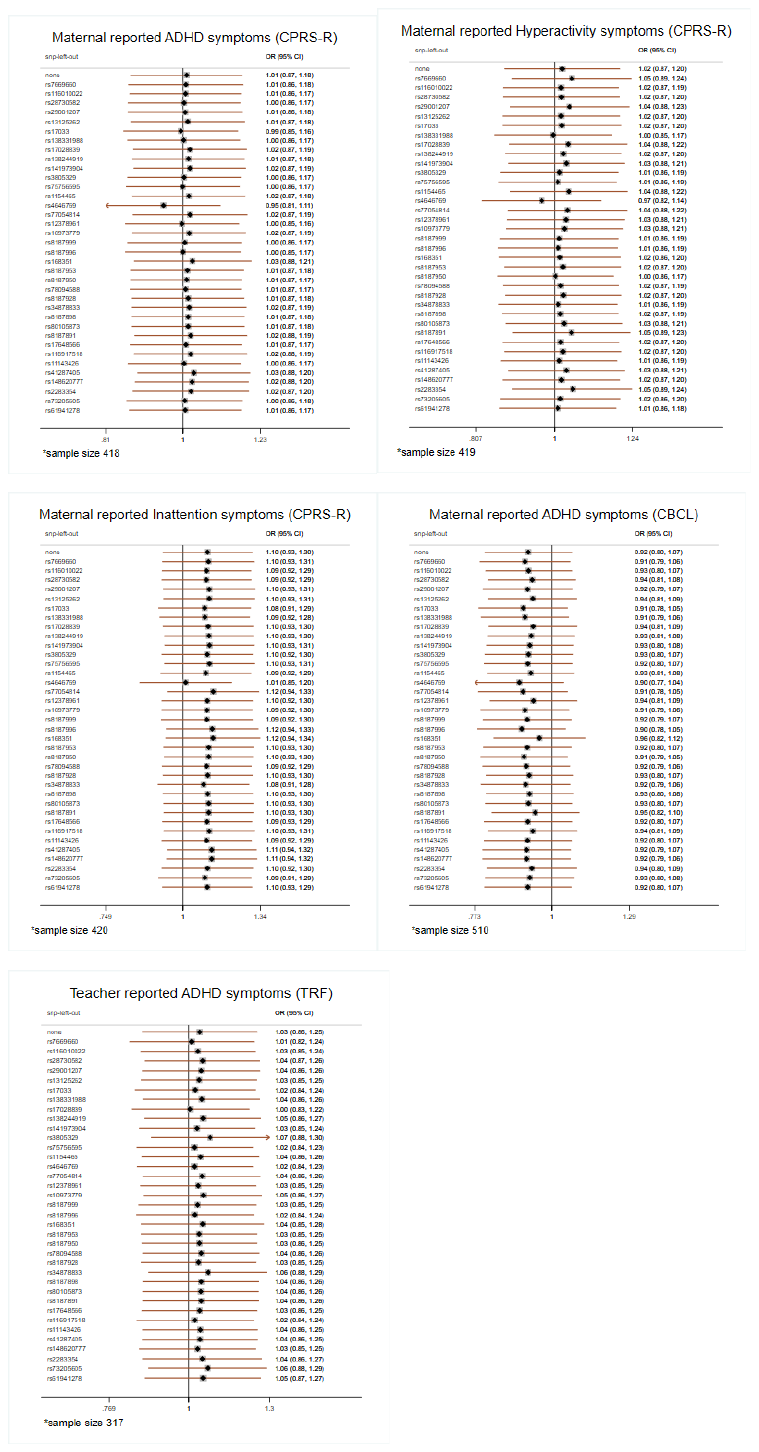

*Note: Revised Conner’s Parent Rating Scale (CGRS-R); Child Behaviour Checklist (CBCL); Teacher Report Form (TRF)*

### Supplementary Figure S13. Leave-one-out analyses in MoBa with maternal GRS on high risk of maternal reported ADHD symptoms

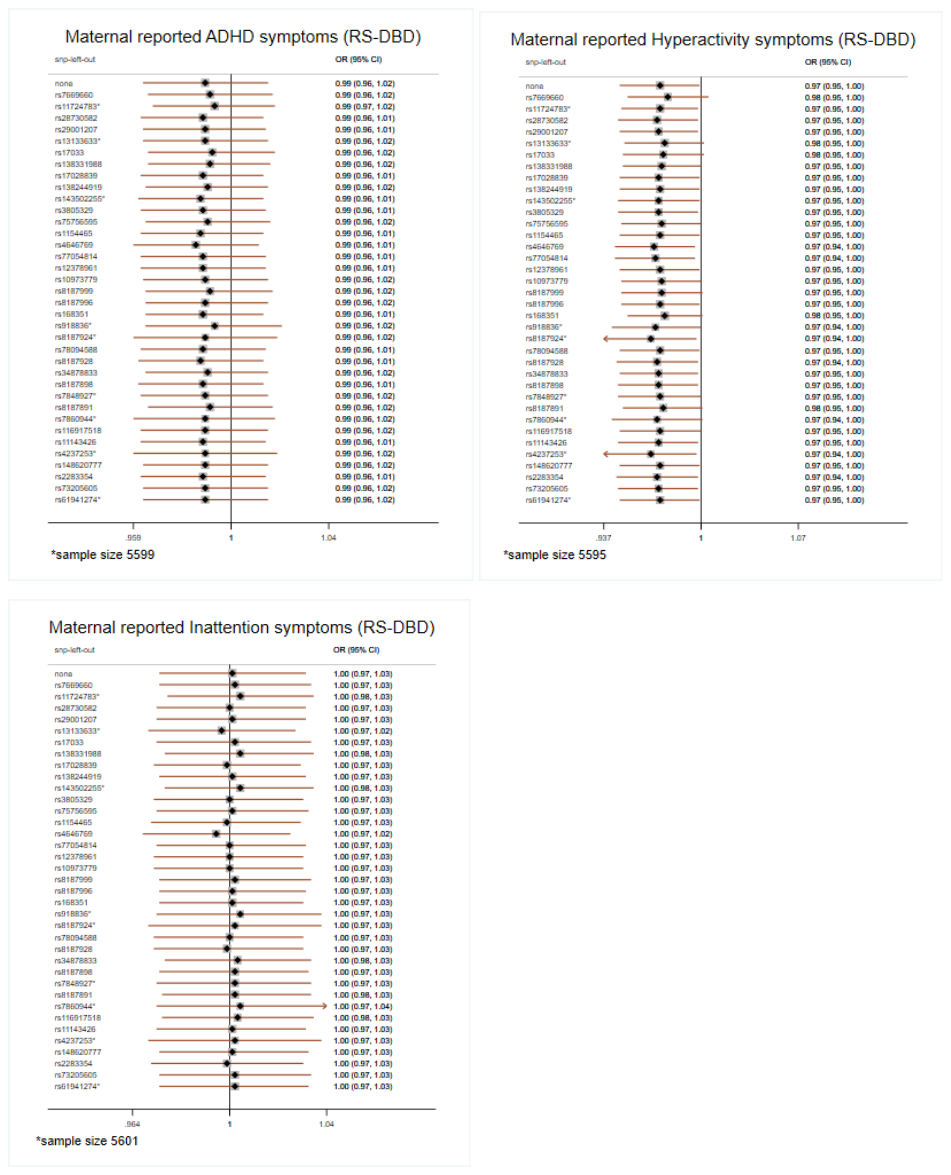

*Note: *new proxy SNP; Disruptive Behaviour Disorders scale (RS-DBD)*

### Supplementary Figure S14. Leave-one-out analyses in MoBa with offspring GRS on high risk of maternal reported ADHD symptoms if mother drink during pregnancy

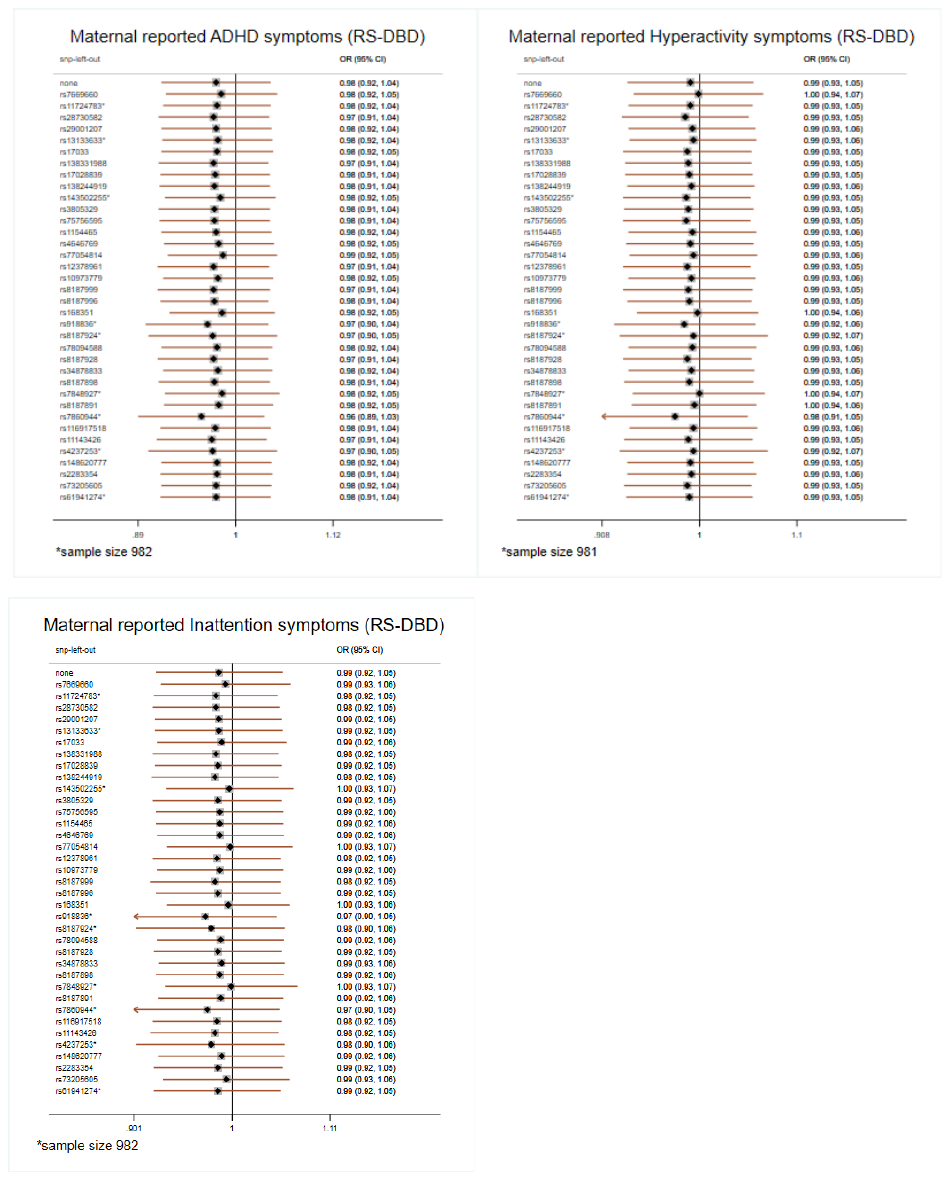

*Note: *new proxy SNP; Disruptive Behaviour Disorders scale (RS-DBD)*

### Supplementary Figure S15. Leave-one-out analyses in MoBa with offspring GRS on high risk of maternal reported ADHD symptoms if mother did not drink during pregnancy

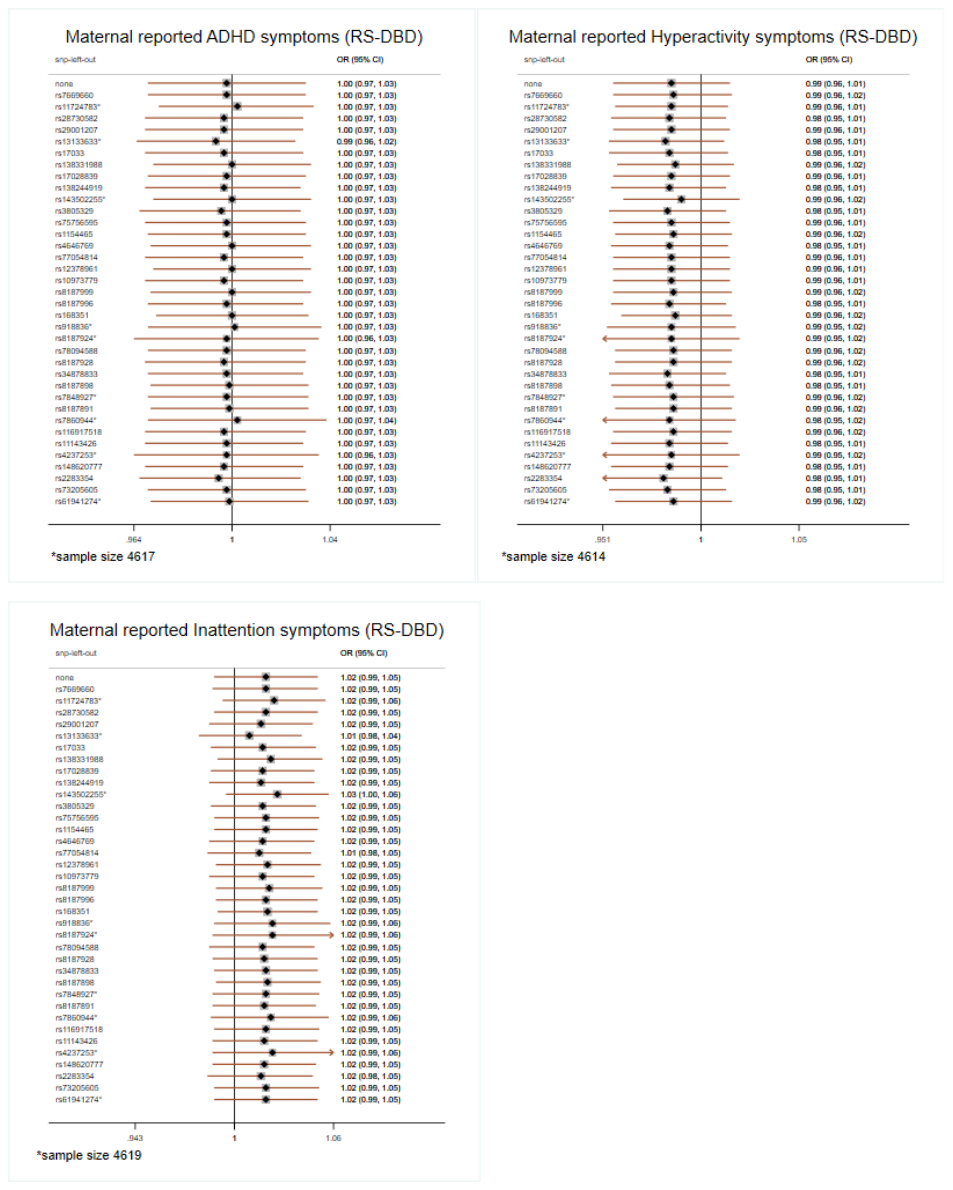

*Note: *new proxy SNP; Disruptive Behaviour Disorders scale (RS-DBD)*

### Supplementary Table S7. Associations between maternal GRS and high risk of maternal reported offspring ADHD symptoms in MoBa

|  | **Model 1** | | | | **Model 2** | | | | **Model 3** | | | | |
| --- | --- | --- | --- | --- | --- | --- | --- | --- | --- | --- | --- | --- | --- |
| **Outcome** | **OR** | **95% CI** | | **P-value** | **OR** | **95% CI** | | **P-value** | **OR** | **95% CI** | | **P-value** | **Sample size** |
| ADHD symptoms (RS-DBD) | 0.99 | 0.963 | 1.016 | 0.418 | 0.99 | 0.959 | 1.020 | 0.488 | 0.99 | 0.956 | 1.022 | 0.494 | 5,599 |
| Hyperactivity symptoms | 0.97 | 0.947 | 0.999 | 0.042 | 0.97 | 0.943 | 1.004 | 0.082 | 0.97 | 0.934 | 0.998 | 0.039 | 5,595 |
| Inattention symptoms | 1.00 | 0.974 | 1.029 | 0.940 | 0.99 | 0.961 | 1.026 | 0.659 | 1.00 | 0.961 | 1.031 | 0.798 | 5,601 |

*Note: Model 1 – only maternal GRS; Model 2 – maternal GRS adj. for offspring GRS; Model 3 – maternal GRS adj. for offspring and paternal GRS; all analyses adjusted for 10 ancestry principal components, birth year and genotyping batch; Disruptive Behaviour Disorders scale (RS-DBD)*

### Supplementary Figure S16. Associations between maternal GRS and high risk of teacher

### reported offspring ADHD symptoms in ALSPAC

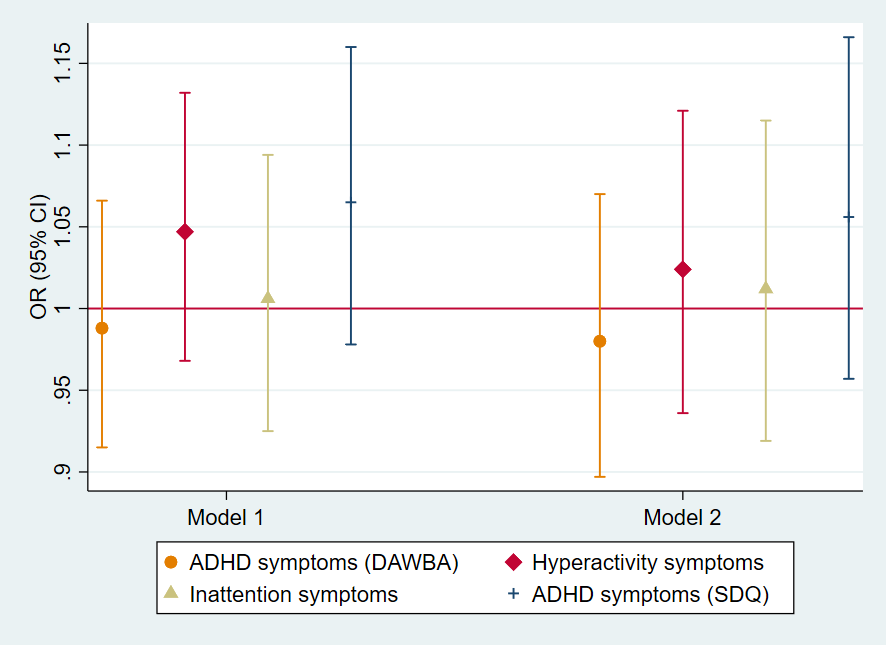

*Note: Model 1 – only maternal GRS; Model 2 – maternal GRS adjusted for offspring GRS; all analyses*

*adjusted for 10 ancestry principal components*; *Development And Well-Being Assessment (DAWBA);*

*Strength and Difficulties Questionnaire (SDQ) – secondary measure*

### Supplementary Table S8. Associations between maternal GRS and high risk of maternal reported offspring ADHD symptoms in ALSPAC

|  | **Model 1** | | | | **Model 2** | | | | |
| --- | --- | --- | --- | --- | --- | --- | --- | --- | --- |
| **Outcome** | **OR** | **95% CI** | | **P-value** | **OR** | **95% CI** | | **P-value** | **Sample size** |
| ADHD symptoms (DAWBA) | 0.99 | 0.938 | 1.047 | 0.746 | 1.01 | 0.951 | 1.080 | 0.682 | 3,698 |
| Hyperactivity symptoms | 1.01 | 0.950 | 1.066 | 0.827 | 1.02 | 0.952 | 1.088 | 0.608 | 3,706 |
| Inattention symptoms | 0.98 | 0.926 | 1.035 | 0.452 | 1.01 | 0.942 | 1.072 | 0.872 | 3,708 |
| ADHD symptoms (SDQ) | 0.96 | 0.897 | 1.017 | 0.148 | 0.95 | 0.883 | 1.020 | 0.159 | 3,736 |

*Note: Model 1 – only maternal GRS; Model 2 – maternal GRS adj. for offspring GRS; all analyses adjusted for 10 ancestry principal components; Development*

*And Well-Being Assessment (DAWBA); Strength and Difficulties Questionnaire (SDQ) – secondary measure*

### Supplementary Table S9. Associations between maternal GRS and high risk of teacher reported offspring ADHD symptoms in ALSPAC

|  | **Model 1** | | | | **Model 2** | | | | |
| --- | --- | --- | --- | --- | --- | --- | --- | --- | --- |
| **Outcome** | **OR** | **95% CI** | | **P-value** | **OR** | **95% CI** | | **P-value** | **Sample size** |
| ADHD symptoms (DAWBA) | 0.99 | 0.915 | 1.066 | 0.750 | 0.98 | 0.897 | 1.070 | 0.649 | 2,310 |
| Hyperactivity symptoms | 1.05 | 0.968 | 1.132 | 0.252 | 1.02 | 0.936 | 1.121 | 0.599 | 2,309 |
| Inattention symptoms | 1.01 | 0.925 | 1.094 | 0.889 | 1.01 | 0.919 | 1.115 | 0.803 | 2,311 |
| ADHD symptoms (SDQ) | 1.07 | 0.978 | 1.160 | 0.148 | 1.06 | 0.957 | 1.166 | 0.277 | 2,312 |

*Note: Model 1 – only maternal GRS; Model 2 – maternal GRS adj. for offspring GRS; all analyses adjusted for 10 ancestry principal components; Development*

*And Well-Being Assessment (DAWBA); Strength and Difficulties Questionnaire (SDQ)*

### Supplementary Table S10. Associations between offspring GRS and high risk of maternal reported offspring ADHD symptoms in ALSPAC

### if mother drink during pregnancy

|  | **Model 1** | | | | **Model 2** | | | | |
| --- | --- | --- | --- | --- | --- | --- | --- | --- | --- |
| **Outcome** | **OR** | **95% CI** | | **P-value** | **OR** | **95% CI** | | **P-value** | **Sample size** |
| ADHD symptoms (DAWBA) | 0.97 | 0.907 | 1.034 | 0.341 | 0.97 | 0.897 | 1.045 | 0.400 | 2,573 |
| Hyperactivity symptoms | 0.98 | 0.913 | 1.050 | 0.552 | 0.98 | 0.903 | 1.061 | 0.599 | 2,576 |
| Inattention symptoms | 0.96 | 0.901 | 1.029 | 0.266 | 0.97 | 0.898 | 1.047 | 0.431 | 2,582 |
| ADHD symptoms (SDQ) | 0.98 | 0.912 | 1.058 | 0.643 | 0.99 | 0.907 | 1.077 | 0.787 | 2,606 |

*Note: Model 1 – only offspring GRS; Model 2 – offspring GRS adj. for maternal GRS; all analyses adjusted for 10 ancestry principal components; Development*

*And Well-Being Assessment (DAWBA); Strength and Difficulties Questionnaire (SDQ) – secondary measure*

### Supplementary Table S11. Associations between offspring GRS and high risk of maternal reported offspring ADHD symptoms in ALSPAC

### if mother did not drink during pregnancy

|  | **Model 1** | | | | **Model 2** | | | | |
| --- | --- | --- | --- | --- | --- | --- | --- | --- | --- |
| **Outcome** | **OR** | **95% CI** | | **P-value** | **OR** | **95% CI** | | **P-value** | **Sample size** |
| ADHD symptoms (DAWBA) | 0.93 | 0.833 | 1.027 | 0.145 | 0.91 | 0.809 | 1.031 | 0.142 | 1,125 |
| Hyperactivity symptoms | 0.97 | 0.875 | 1.085 | 0.635 | 0.96 | 0.847 | 1.085 | 0.502 | 1,130 |
| Inattention symptoms | 0.90 | 0.808 | 1.002 | 0.055 | 0.89 | 0.788 | 1.009 | 0.069 | 1,126 |
| ADHD symptoms (SDQ) | 0.99 | 0.878 | 1.116 | 0.868 | 1.07 | 0.929 | 1.227 | 0.356 | 1,130 |

*Note: Model 1 – only offspring GRS; Model 2 – offspring GRS adj. for maternal GRS; all analyses adjusted for 10 ancestry principal components; Development*

*And Well-Being Assessment (DAWBA); Strength and Difficulties Questionnaire (SDQ) – secondary measure*

### Supplementary Table S12. Associations between offspring GRS and high risk of maternal reported offspring ADHD symptoms in MoBa if mother drink during pregnancy

|  | **Model 1** | | | | **Model 2** | | | | **Model 3** | | | | |
| --- | --- | --- | --- | --- | --- | --- | --- | --- | --- | --- | --- | --- | --- |
| **Outcome** | **OR** | **95% CI** | | **P-value** | **OR** | **95% CI** | | **P-value** | **OR** | **95% CI** | | **P-value** | **Sample size** |
| ADHD symptoms (RS-DBD) | 0.98 | 0.915 | 1.043 | 0.485 | 0.99 | 0.917 | 1.073 | 0.842 | 0.98 | 0.895 | 1.081 | 0.728 | 982 |
| Hyperactivity symptoms | 0.99 | 0.931 | 1.055 | 0.778 | 1.02 | 0.944 | 1.097 | 0.656 | 1.02 | 0.933 | 1.115 | 0.666 | 981 |
| Inattention symptoms | 0.99 | 0.922 | 1.055 | 0.692 | 0.98 | 0.898 | 1.059 | 0.548 | 0.96 | 0.869 | 1.062 | 0.431 | 982 |

*Note: Model 1 – only offspring GRS; Model 2 – offspring GRS adj. for maternal GRS; Model 3 – offspring GRS adj. for maternal and paternal GRS; all analyses adjusted for 10 ancestry principal components, birth year and genotyping batch; Disruptive Behaviour Disorders scale (RS-DBD)*

### Supplementary Table S13. Associations between offspring GRS and high risk of maternal reported offspring ADHD symptoms in MoBa if mother did not drink during pregnancy

|  | **Model 1** | | | | **Model 2** | | | | **Model 3** | | | | |
| --- | --- | --- | --- | --- | --- | --- | --- | --- | --- | --- | --- | --- | --- |
| **Outcome** | **OR** | **95% CI** | | **P-value** | **OR** | **95% CI** | | **P-value** | **OR** | **95% CI** | | **P-value** | **Sample size** |
| ADHD symptoms (RS-DBD) | 1.00 | 0.969 | 1.028 | 0.904 | 1.00 | 0.968 | 1.038 | 0.881 | 1.01 | 0.965 | 1.050 | 0.766 | 4,617 |
| Hyperactivity symptoms | 0.99 | 0.956 | 1.015 | 0.318 | 1.00 | 0.965 | 1.035 | 0.986 | 1.02 | 0.973 | 1.061 | 0.479 | 4,614 |
| Inattention symptoms | 1.02 | 0.988 | 1.051 | 0.242 | 1.03 | 0.988 | 1.063 | 0.193 | 1.02 | 0.977 | 1.069 | 0.353 | 4,619 |

*Note: Model 1 – only offspring GRS; Model 2 – offspring GRS adj. for maternal GRS; Model 3 – offspring GRS adj. for maternal and paternal GRS; all analyses adjusted for 10 ancestry principal components, birth year and genotyping batch; Disruptive Behaviour Disorders scale (RS-DBD)*

### Supplementary Figure S17. Associations between offspring GRS and high risk of teacher reported offspring ADHD symptoms in ALSPAC stratified by maternal drinking status

**Mother did not drink during pregnancy**

**Mother drink during pregnancy**

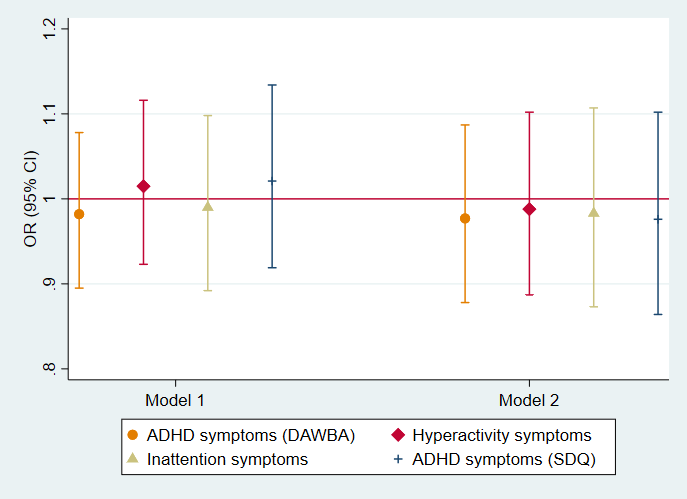

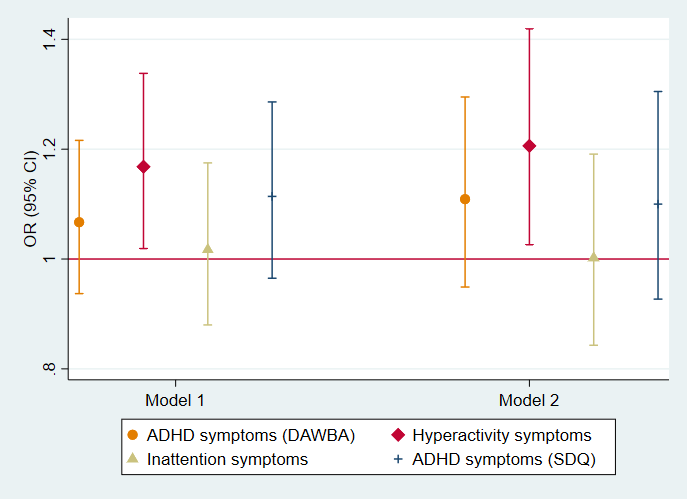

*Note: Model 1 – only offspring GRS; Model 2 – offspring GRS adjusted for maternal GRS; all analyses adjusted for 10 ancestry principal components*; *Development And Well-Being Assessment (DAWBA); Strength and Difficulties Questionnaire (SDQ)*

### Supplementary Table S14. Associations between offspring GRS and high risk of maternal and teacher reported offspring ADHD symptoms in GenR

|  | **Mother drink during pregnancy** | | | | | **Mother did not drink during pregnancy** | | | | |
| --- | --- | --- | --- | --- | --- | --- | --- | --- | --- | --- |
| **Outcome** | **OR** | **95% CI** | | **P-value** | **Sample size** | **OR** | **95% CI** | | **P-value** | **Sample size** |
| ADHD symptoms (CGRS-R) | 0.99 | 0.895 | 1.083 | 0.750 | 1037 | 1.01 | 0.867 | 1.176 | 0.903 | 418 |
| Hyperactivity symptoms | 1.00 | 0.896 | 1.123 | 0.958 | 1038 | 1.02 | 0.872 | 1.198 | 0.790 | 419 |
| Inattention symptoms | 0.99 | 0.891 | 1.093 | 0.794 | 1038 | 1.10 | 0.927 | 1.296 | 0.285 | 420 |
| ADHD symptoms (CBCL) | 1.04 | 0.945 | 1.152 | 0.398 | 1186 | 0.92 | 0.799 | 1.069 | 0.288 | 510 |
| ADHD symptoms (TRF) | 1.00 | 0.860 | 1.157 | 0.976 | 708 | 1.04 | 0.857 | 1.251 | 0.719 | 317 |

*Note: Model 1 – only offspring GRS adjusted for 10 ancestry principal components; Revised Conner’s Parent Rating Scale (CGRS-R); Child Behaviour Checklist (CBCL); Teacher Report Form (TRF); CBCL and TRF are secondary measures*

### Supplementary Table S15. Associations between offspring GRS and high risk of teacher reported offspring ADHD symptoms in ALSPAC

### if mother drink during pregnancy

|  | **Model 1** | | | | **Model 2** | | | | |
| --- | --- | --- | --- | --- | --- | --- | --- | --- | --- |
| **Outcome** | **OR** | **95% CI** | | **P-value** | **OR** | **95% CI** | | **P-value** | **Sample size** |
| ADHD symptoms (DAWBA) | 0.98 | 0.895 | 1.078 | 0.706 | 0.98 | 0.878 | 1.087 | 0.666 | 1,602 |
| Hyperactivity symptoms | 1.02 | 0.923 | 1.116 | 0.755 | 0.99 | 0.887 | 1.102 | 0.832 | 1,601 |
| Inattention symptoms | 0.99 | 0.892 | 1.098 | 0.843 | 0.98 | 0.873 | 1.107 | 0.780 | 1,603 |
| ADHD symptoms (SDQ) | 1.02 | 0.919 | 1.134 | 0.701 | 0.98 | 0.864 | 1.102 | 0.691 | 1,603 |

*Note: Model 1 – only offspring GRS; Model 2 – offspring GRS adj. for maternal GRS; all analyses adjusted for 10 ancestry principal components; Development*

*And Well-Being Assessment (DAWBA); Strength and Difficulties Questionnaire (SDQ)*

### Supplementary Table S16. Associations between offspring GRS and high risk of teacher reported offspring ADHD symptoms in ALSPAC

### if mother did not drink during pregnancy

|  | **Model 1** | | | | **Model 2** | | | | |
| --- | --- | --- | --- | --- | --- | --- | --- | --- | --- |
| **Outcome** | **OR** | **95% CI** | | **P-value** | **OR** | **95% CI** | | **P-value** | **Sample size** |
| ADHD symptoms (DAWBA) | 1.07 | 0.937 | 1.216 | 0.329 | 1.11 | 0.949 | 1.295 | 0.192 | 708 |
| Hyperactivity symptoms | 1.17 | 1.019 | 1.338 | 0.025 | 1.21 | 1.026 | 1.419 | 0.023 | 708 |
| Inattention symptoms | 1.02 | 0.880 | 1.175 | 0.822 | 1.00 | 0.843 | 1.191 | 0.985 | 708 |
| ADHD symptoms (SDQ) | 1.11 | 0.965 | 1.286 | 0.142 | 1.10 | 0.927 | 1.305 | 0.277 | 709 |

*Note: Model 1 – only offspring GRS; Model 2 – offspring GRS adj. for maternal GRS; all analyses adjusted for 10 ancestry principal components; Development*

*And Well-Being Assessment (DAWBA); Strength and Difficulties Questionnaire (SDQ)*

### Supplementary Table S17. Associations between maternal GRS and confounders in ALSPAC

| **Confounder** | **Effect estimate** | **Effect size** | **95% CI** | **P-value** | **Sample size** |
| --- | --- | --- | --- | --- | --- |
| Maternal age | Beta | -0.02 | -0.084, 0.042 | 0.507 | 7,421 |
| Maternal education | Beta | 0.001 | -0.017, 0.018 | 0.951 | 6,860 |
| Financial difficulties | Beta | -0.03 | -0.078, 0.018 | 0.217 | 6,691 |
| Marital status | OR | 0.98 | 0.949, 1.013 | 0.227 | 7,124 |
| Depression symptoms | OR | 1.02 | 0.982, 1.069 | 0.264 | 6,706 |
| Anxiety symptoms | OR | 0.98 | 0.945, 1.022 | 0.384 | 6,669 |
| Parity | Beta | 0.01 | -0.004, 0.020 | 0.184 | 7,040 |
| Maternal smoking in pregnancy | OR | 0.90 | 0.958,1.022 | 0.528 | 7,138 |

*Note: adjusted for 10 ancestry principal components; OR – odds ratio; 95% CI – 95% confidence intervals.*

### Supplementary Table S18. Associations between maternal GRS and confounders in MoBa

| **Confounder** | **Effect estimate** | **Effect size** | **95% CI** | **P-value** | **Sample size** |
| --- | --- | --- | --- | --- | --- |
| Maternal age | Beta | 0.04 | -0.049, 0.129 | 0.381 | 13,614 |
| Maternal education | Beta | 0.0003 | -0.003, 0.0003 | 0.839 | 12,917 |
| Financial difficulties | OR | 1.00 | 0.980, 1.012 | 0.650 | 12,575 |
| Marital status | Beta | 0.001 | 0.0001, 0.002 | 0.035 | 13,555 |
| Depression/Anxiety symptoms | OR | 0.99 | 0.969, 1.014 | 0.454 | 13,503 |
| Parity | Beta | 0.002 | -0.003, 0.007 | 0.433 | 13,614 |
| Maternal ADHD symptoms | OR | 1.02 | 0.976,1.065 | 0.390 | 8,231 |
| Maternal smoking in pregnancy | OR | 1.00 | 0.973, 1.016 | 0.627 | 13,540 |

*Note: adjusted for 10 ancestry principal components; birth year and genotyping batch; OR – odds ratio; 95% CI – 95% confidence intervals*

### Supplementary Figure S18. Associations between offspring GRS and high risk of

### maternal and teacher reported offspring ADHD symptoms in GenR stratified by

### maternal drinking status

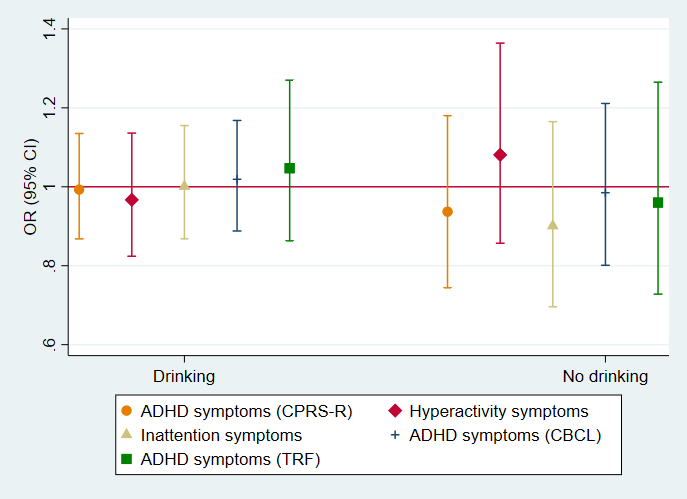

*Note: Model 1 – only offspring GRS adjusted for 10 ancestry principal components; offspring GRS*

*of 4 SNPs (rs2866151, rs975833, rs4147536, rs284779); Revised Conner’s Parent Rating Scale*

*(CPRS-R); Child Behaviour Checklist (CBCL); Teacher Report Form (TRF); CBCL and TRF are secondary*

*measures*

### Supplementary Figure S19. Associations between offspring GRS and high risk of maternal reported offspring ADHD symptoms in ALSPAC stratified by maternal drinking status

**Mother did not drink during pregnancy**

**Mother drink during pregnancy**

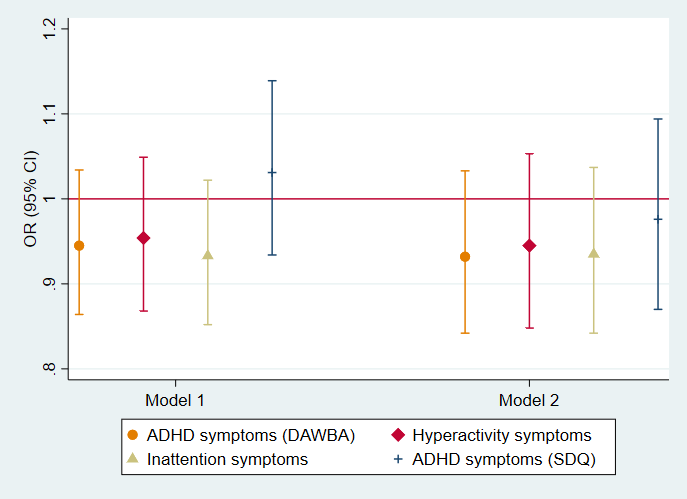

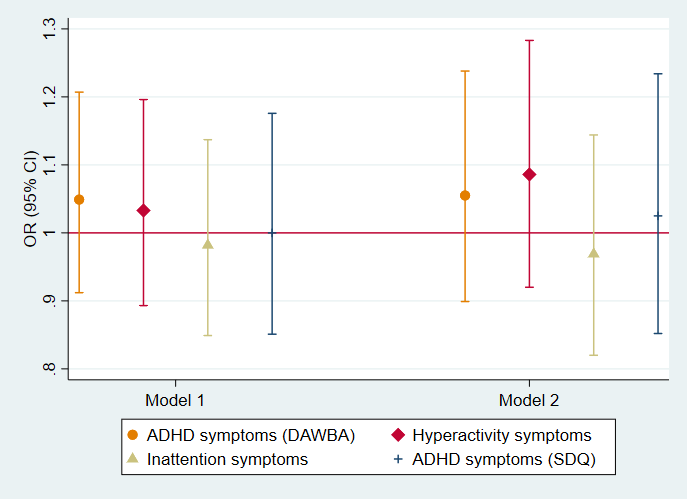

*Note: Model 1 – only offspring GRS; Model 2 – offspring GRS adjusted for maternal GRS; all analyses adjusted for 10 ancestry principal components*; *offspring GRS of 4 SNPs (rs2866151, rs975833, rs4147536, rs284779); Development And Well-Being Assessment (DAWBA); Strength and Difficulties Questionnaire (SDQ) – secondary measure*

### Supplementary Figure S20. Associations between offspring GRS and high risk of teacher reported offspring ADHD symptoms in ALSPAC stratified by maternal drinking status

**Mother did not drink during pregnancy**

**Mother drink during pregnancy**

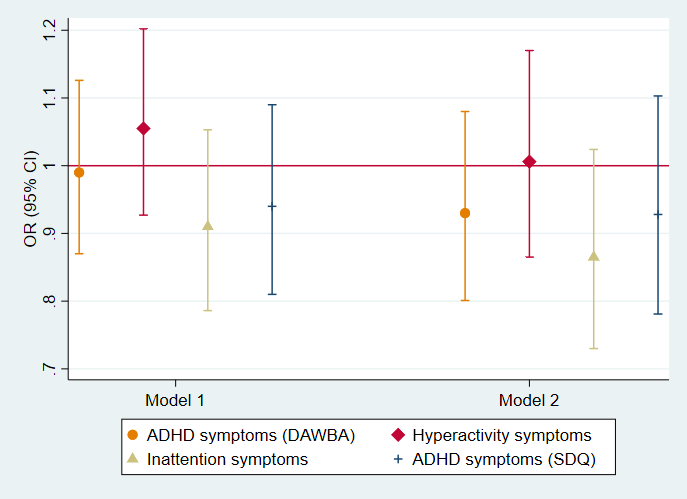

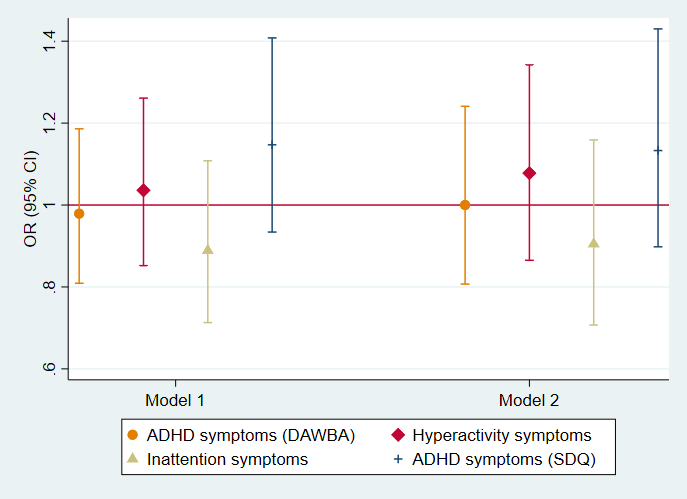

*Note: Model 1 – only offspring GRS; Model 2 – offspring GRS adjusted for maternal GRS; all analyses adjusted for 10 ancestry principal components*; *offspring GRS of 4 SNPs (rs2866151, rs975833, rs4147536, rs284779); Development And Well-Being Assessment (DAWBA); Strength and Difficulties Questionnaire (SDQ)*

### Supplementary Figure S21. Associations between offspring GRS and high risk of maternal reported offspring ADHD symptoms in MoBa stratified by maternal drinking status

**Mother drink during pregnancy**

**Mother did not drink during pregnancy**

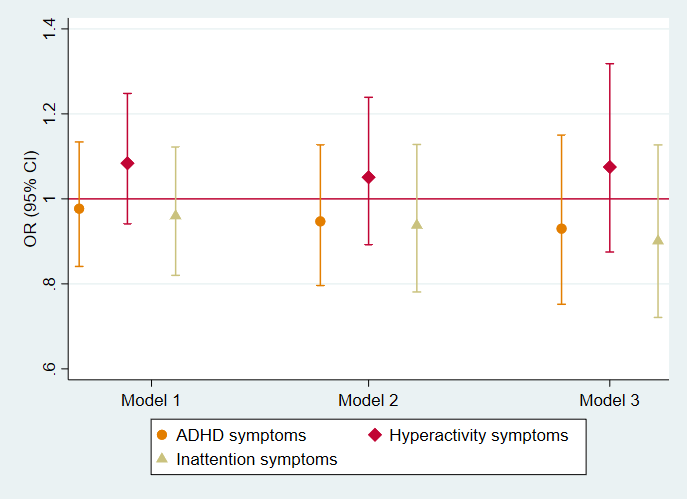

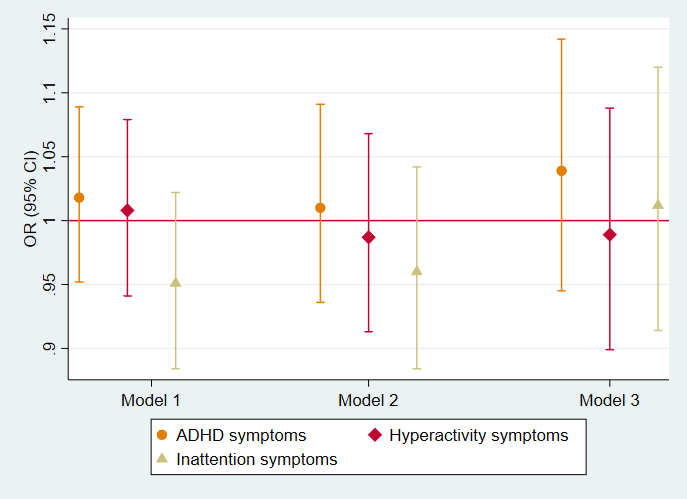

*Note: Model 1 – only offspring GRS; Model 2 – offspring GRS adjusted for maternal GRS; Model 3 – offspring GRS adjusted for maternal and paternal GRS; all analyses are adjusted for 10 ancestry principal components, birth year and genotyping batch; offspring GRS of 4 SNPs (rs2866151, rs975833, rs4147536, rs284779);*

*ADHD symptoms measured with Disruptive Behaviour Disorders Scale (RS-DBD)*

### Supplementary Table S19. Associations between offspring GRS and high risk of maternal and teacher reported offspring ADHD symptoms in GenR

|  | **Mother drink during pregnancy** | | | | | **Mother did not drink during pregnancy** | | | | |
| --- | --- | --- | --- | --- | --- | --- | --- | --- | --- | --- |
| **Outcome** | **OR** | **95% CI** | | **P-value** | **Sample size** | **OR** | **95% CI** | | **P-value** | **Sample size** |
| ADHD symptoms (CGRS-R) | 0.99 | 0.868 | 1.135 | 0.915 | 1037 | 0.94 | 0.744 | 1.180 | 0.581 | 418 |
| Hyperactivity symptoms | 0.97 | 0.824 | 1.136 | 0.687 | 1038 | 1.08 | 0.857 | 1.364 | 0.512 | 419 |
| Inattention symptoms | 1.00 | 0.868 | 1.155 | 0.990 | 1038 | 0.90 | 0.696 | 1.165 | 0.426 | 420 |
| ADHD symptoms (CBCL) | 1.02 | 0.888 | 1.168 | 0.793 | 1186 | 0.99 | 0.801 | 1.211 | 0.884 | 510 |
| ADHD symptoms (TRF) | 1.05 | 0.863 | 1.270 | 0.639 | 708 | 0.96 | 0.728 | 1.265 | 0.770 | 317 |

*Note: Model 1 – only offspring GRS adjusted also for 10 ancestry principal components; offspring GRS of 4 SNPs (rs2866151, rs975833, rs4147536, rs284779);*

*Revised Conner’s Parent Rating Scale (CGRS-R); Child Behaviour Checklist (CBCL); Teacher Report Form (TRF); CBCL and TRF are secondary measures*

### Supplementary Table S20. Associations between offspring GRS and high risk of maternal reported offspring ADHD symptoms in ALSPAC

### if mother drink during pregnancy

|  | **Model 1** | | | | **Model 2** | | | | |
| --- | --- | --- | --- | --- | --- | --- | --- | --- | --- |
| **Outcome** | **OR** | **95% CI** | | **P-value** | **OR** | **95% CI** | | **P-value** | **Sample size** |
| ADHD symptoms (DAWBA) | 0.95 | 0.864 | 1.034 | 0.218 | 0.93 | 0.842 | 1.033 | 0.181 | 2,573 |
| Hyperactivity symptoms | 0.95 | 0.868 | 1.049 | 0.328 | 0.95 | 0.848 | 1.053 | 0.307 | 2,576 |
| Inattention symptoms | 0.93 | 0.852 | 1.022 | 0.134 | 0.94 | 0.842 | 1.037 | 0.201 | 2,582 |
| ADHD symptoms (SDQ) | 1.03 | 0.934 | 1.139 | 0.544 | 0.98 | 0.870 | 1.094 | 0.672 | 2,606 |

*Note: Model 1 – only offspring GRS; Model 2 – offspring GRS adj. for maternal GRS; all analyses adjusted for 10 ancestry principal components; offspring*

*GRS of 4 SNPs (rs2866151, rs975833, rs4147536, rs284779); Development And Well-Being Assessment (DAWBA); Strength and Difficulties Questionnaire*

*(SDQ) – secondary measure*

### Supplementary Table S21. Associations between offspring GRS and high risk of maternal reported offspring ADHD symptoms in ALSPAC

### if mother did not drink during pregnancy

|  | **Model 1** | | | | **Model 2** | | | | |
| --- | --- | --- | --- | --- | --- | --- | --- | --- | --- |
| **Outcome** | **OR** | **95% CI** | | **P-value** | **OR** | **95% CI** | | **P-value** | **Sample size** |
| ADHD symptoms (DAWBA) | 1.05 | 0.912 | 1.207 | 0.504 | 1.06 | 0.899 | 1.238 | 0.514 | 1,125 |
| Hyperactivity symptoms | 1.03 | 0.893 | 1.196 | 0.660 | 1.09 | 0.920 | 1.283 | 0.330 | 1,130 |
| Inattention symptoms | 0.98 | 0.849 | 1.137 | 0.808 | 0.97 | 0.820 | 1.144 | 0.708 | 1,126 |
| ADHD symptoms (SDQ) | 1.00 | 0.851 | 1.1758 | 0.996 | 1.03 | 0.852 | 1.234 | 0.790 | 1,130 |

*Note: Model 1 – only offspring GRS; Model 2 – offspring GRS adj. for maternal GRS; all analyses adjusted for 10 ancestry principal components; offspring*

*GRS of 4 SNPs (rs2866151, rs975833, rs4147536, rs284779); Development And Well-Being Assessment (DAWBA); Strength and Difficulties Questionnaire*

*(SDQ) – secondary measure*

### Supplementary Table S22. Associations between offspring GRS and high risk of teacher reported ADHD symptoms in ALSPAC if mother

### drink during pregnancy

|  | **Model 1** | | | | **Model 2** | | | | |
| --- | --- | --- | --- | --- | --- | --- | --- | --- | --- |
| **Outcome** | **OR** | **95% CI** | | **P-value** | **OR** | **95% CI** | | **P-value** | **Sample size** |
| ADHD symptoms (DAWBA) | 0.99 | 0.870 | 1.126 | 0.876 | 0.93 | 0.801 | 1.080 | 0.344 | 1,602 |
| Hyperactivity symptoms | 1.06 | 0.927 | 1.202 | 0.416 | 1.01 | 0.865 | 1.170 | 0.940 | 1,601 |
| Inattention symptoms | 0.91 | 0.786 | 1.053 | 0.206 | 0.87 | 0.730 | 1.024 | 0.092 | 1,603 |
| ADHD symptoms (SDQ) | 0.94 | 0.810 | 1.090 | 0.413 | 0.93 | 0.781 | 1.103 | 0.398 | 1,603 |

*Note: Model 1 – only offspring GRS; Model 2 – offspring GRS adj. for maternal GRS; all analyses adjusted for 10 ancestry principal components; offspring GRS*

*of 4 SNPs (rs2866151, rs975833, rs4147536, rs284779); Development And Well-Being Assessment (DAWBA); Strength and Difficulties Questionnaire (SDQ)*

### Supplementary Table S23. Associations between offspring GRS and high risk of teacher reported offspring ADHD symptoms in ALSPAC

### if mother did not drink during pregnancy

|  | **Model 1** | | | | **Model 2** | | | | |
| --- | --- | --- | --- | --- | --- | --- | --- | --- | --- |
| **Outcome** | **OR** | **95% CI** | | **P-value** | **OR** | **95% CI** | | **P-value** | **Sample size** |
| ADHD symptoms (DAWBA) | 0.98 | 0.809 | 1.186 | 0.830 | 1.00 | 0.807 | 1.241 | 0.998 | 708 |
| Hyperactivity symptoms | 1.04 | 0.852 | 1.261 | 0.723 | 1.08 | 0.865 | 1.342 | 0.504 | 708 |
| Inattention symptoms | 0.89 | 0.713 | 1.108 | 0.293 | 0.91 | 0.707 | 1.159 | 0.429 | 708 |
| ADHD symptoms (SDQ) | 1.15 | 0.934 | 1.408 | 0.191 | 1.13 | 0.898 | 1.430 | 0.292 | 709 |

*Note: Model 1 – only offspring GRS; Model 2 – offspring GRS adj. for maternal GRS; all analyses adjusted also for 10 ancestry principal components; offspring*

*GRS of 4 SNPs (rs2866151, rs975833, rs4147536, rs284779); Development And Well-Being Assessment (DAWBA); Strength and Difficulties Questionnaire (SDQ)*

### Supplementary Table S24. Associations between offspring GRS and high risk of maternal reported offspring ADHD symptoms in MoBa if mother drink during pregnancy

|  | **Model 1** | | | | **Model 2** | | | | **Model 3** | | | | |
| --- | --- | --- | --- | --- | --- | --- | --- | --- | --- | --- | --- | --- | --- |
| **Outcome** | **OR** | **95% CI** | | **P-value** | **OR** | **95% CI** | | **P-value** | **OR** | **95% CI** | | **P-value** | **Sample size** |
| ADHD symptoms (RS-DBD) | 0.98 | 0.841 | 1.134 | 0.757 | 0.95 | 0.796 | 1.127 | 0.538 | 0.93 | 0.752 | 1.150 | 0.503 | 982 |
| Hyperactivity symptoms | 1.08 | 0.941 | 1.248 | 0.263 | 1.05 | 0.892 | 1.239 | 0.553 | 1.08 | 0.875 | 1.318 | 0.494 | 981 |
| Inattention symptoms | 0.96 | 0.820 | 1.122 | 0.606 | 0.94 | 0.781 | 1.128 | 0.498 | 0.90 | 0.721 | 1.127 | 0.362 | 982 |

*Note: Model 1 – only offspring GRS; Model 2 – offspring GRS adj. for maternal GRS; Model 3 – offspring GRS adj. for maternal and paternal GRS; all analyses adjusted for 10 ancestry principal components, birth year and genotyping batch; offspring GRS of 4 SNPs (rs2866151, rs975833, rs4147536, rs284779); Disruptive Behaviour Disorders scale (RS-DBD)*

### Supplementary Table S25. Associations between offspring GRS and high risk of maternal reported offspring ADHD symptoms in MoBa if mother did not drink during pregnancy

|  | **Model 1** | | | | **Model 2** | | | | **Model 3** | | | | |
| --- | --- | --- | --- | --- | --- | --- | --- | --- | --- | --- | --- | --- | --- |
| **Outcome** | **OR** | **95% CI** | | **P-value** | **OR** | **95% CI** | | **P-value** | **OR** | **95% CI** | | **P-value** | **Sample size** |
| ADHD symptoms (RS-DBD) | 1.02 | 0.952 | 1.089 | 0.605 | 1.01 | 0.936 | 1.091 | 0.793 | 1.04 | 0.945 | 1.142 | 0.427 | 4,617 |
| Hyperactivity symptoms | 1.01 | 0.941 | 1.079 | 0.823 | 0.99 | 0.913 | 1.068 | 0.750 | 0.99 | 0.899 | 1.088 | 0.821 | 4,614 |
| Inattention symptoms | 0.95 | 0.884 | 1.022 | 0.168 | 0.96 | 0.884 | 1.042 | 0.329 | 1.01 | 0.914 | 1.120 | 0.820 | 4,619 |

*Note: Model 1 – only offspring GRS; Model 2 – offspring GRS adj. for maternal GRS; Model 3 – offspring GRS adj. for maternal and paternal GRS; all analyses adjusted for 10 ancestry principal components, birth year and genotyping batch; offspring GRS of 4 SNPs (rs2866151, rs975833, rs4147536, rs284779); Disruptive Behaviour Disorders scale (RS-DBD)*

### References

Achenbach, T. M., & Rescorla, L. A. (2001). *Manual for the ASEBA School-Age Forms & Profiles.* Burlington: University of Vermont, Research Centre for Children, Youth and Families.

Alati, R., Smith, G. D., Lewis, S. J., Sayal, K., Draper, E. S., Golding, J., Fraser, R., & Gray, R. (2013). Effect of Prenatal Alcohol Exposure on Childhood Academic Outcomes: Contrasting Maternal and Paternal Associations in the ALSPAC Study. *Plos One, 8*(10). doi:ARTN e7484410.1371/journal.pone.0074844

Bakker, R., Pluimgraaff, L. E., Steegers, E. A. P., Raat, H., Tiemeier, H., Hofman, A., & Jaddoe, V. W. V. (2010). Associations of light and moderate maternal alcohol consumption with fetal growth characteristics in different periods of pregnancy: The Generation R Study. *International Journal of Epidemiology, 39*(3), 777-789. doi:10.1093/ije/dyq047

Conners, C. K., Sitarenios, G., Parker, J. D. A., & Epstein, J. N. (1998). The revised Conners' Parent Rating Scale (CPRS-R): Factor structure, reliability, and criterion validity. *Journal of Abnormal Child Psychology, .26*(4), pp. doi:10.1023/A:1022602400621 9700518

Goodman, R. (1997). The strengths and difficulties questionnaire: A research note. *Journal of Child Psychology and Psychiatry and Allied Disciplines, 38*(5), 581-586. doi:DOI 10.1111/j.1469-7610.1997.tb01545.x

Goodman, R. (2001). Psychometric properties of the strengths and difficulties questionnaire. *Journal of the American Academy of Child and Adolescent Psychiatry, 40*(11), 1337-1345. doi:Doi 10.1097/00004583-200111000-00015

Goodman, R., Ford, T., Richards, H., Gatward, R., & Meltzer, H. (2000). The Development and Well-Being Assessment: Description and initial validation of an integrated assessment of child and adolescent psychopathology. *Journal of Child Psychology and Psychiatry and Allied Disciplines, 41*(5), 645-655. doi:Doi 10.1017/S0021963099005909

Helgeland, O., Vaudel, M., Juliusson, P. B., Holmen, O. L., Juodakis, J., Bacelis, J., Jacobsson, B., Lindekleiv, H., Hveem, K., Lie, R. T., Knudsen, G. P., Stoltenberg, C., Magnus, P., Sagen, J. V., Molven, A., Johansson, S., & Njolstad, P. R. (2019). Genome-wide association study reveals dynamic role of genetic variation in infant and early childhood growth. *Nature Communications, 10*. doi:ARTN 444810.1038/s41467-019-12308-0

Knudsen, A. K., Skogen, J. C., Ystrom, E., Sivertsen, B., Tell, G. S., & Torgersen, L. (2014). Maternal pre-pregnancy risk drinking and toddler behavior problems: the Norwegian Mother and Child Cohort Study. *European Child & Adolescent Psychiatry, 23*(10), 901-911. doi:10.1007/s00787-014-0588-x

Kooijman, M. N., Kruithof, C. J., van Duijn, C. M., Duijts, L., Franco, O. H., van IJzendoorn, M. H., de Jongste, J. C., Klaver, C. C. W., van der Lugt, A., Mackenbach, J. P., Moll, H. A., Peeters, R. P., Raat, H., Rings, E. H. H. M., Rivadeneira, F., van der Schroeff, M. P., Steegers, E. A. P., Tiemeier, H., Uitterlinden, A. G., Verhulst, F. C., Wolvius, E., Felix, J. F., & Jaddoe, V. W. V. (2016). The Generation R Study: design and cohort update 2017. *European Journal of Epidemiology, 31*(12), 1243-1264. doi:10.1007/s10654-016-0224-9

Medina-Gomez, C., Felix, J. F., Estrada, K., Peters, M. J., Herrera, L., Kruithof, C. J., Duijts, L., Hofman, A., van Duijn, C. M., Uitterlinden, A. G., Jaddoe, V. W. V., & Rivadeneira, F. (2015). Challenges in conducting genome-wide association studies in highly admixed multi-ethnic populations: the Generation R Study. *European Journal of Epidemiology, 30*(4), 317-330. doi:10.1007/s10654-015-9998-4

Paternoster, L., Evans, D. M., Nohr, E. A., Holst, C., Gaborieau, V., Brennan, P., Gjesing, A. P., Grarup, N., Witte, D. R., Jorgensen, T., Linneberg, A., Lauritzen, T., Sandbaek, A., Hansen, T., Pedersen, O., Elliott, K. S., Kemp, J. P., St Pourcain, B., McMahon, G., Zelenika, D., Hager, J., Lathrop, M., Timpson, N. J., Smith, G. D., & Sorensen, T. I. A. (2011). Genome-Wide Population-Based Association Study of Extremely Overweight Young Adults - The GOYA Study. *Plos One, 6*(9). doi:ARTN e2430310.1371/journal.pone.0024303

Rishel, C. W., Greeno, C., Marcus, S. C., Shear, M. K., & Anderson, C. (2005). Use of the child behavior checklist as a diagnostic screening tool in community mental health. *Research on Social Work Practice, 15*(3), 195-203. doi:10.1177/1049731504270382

Silva, R. R., Alpert, M., Pouget, E., Silva, V., Trosper, S., Reyes, K., & Dummit, S. (2005). A rating scale for disruptive behavior disorders, based on the DSM-IV item pool. *Psychiatric Quarterly, .76*(4), pp. doi:10.1007/s11126-005-4966-x 16217627

Taylor, A. E., Jones, H. J., Sallis, H., Euesden, J., Stergiakouli, E., Davies, N. M., Zammit, S., Lawlor, D. A., Munafo, M. R., Smith, G. D., & Tilling, K. (2018). Exploring the association of genetic factors with participation in the Avon Longitudinal Study of Parents and Children. *International Journal of Epidemiology, 47*(4), 1207-1216. doi:10.1093/ije/dyy060
